# Multi-ancestry genome-wide and transcriptome-wide association analyses identify new risk loci and genes for inflammatory bowel disease

**DOI:** 10.1101/2025.09.23.25336483

**Authors:** Linshuoshuo Lyu, Qing Li, Chao Li, Ghadeer K Dawwas, You Chen, Ran Tao, Qi Liu, Wanqing Wen, Xiao-Ou Shu, Kay Washington, David A Schwartz, Wei Zheng, Zhijun Yin, Ken S Lau, Xingyi Guo

## Abstract

To advance genetic understanding of inflammatory bowel disease (IBD), we conduct multi-ancestry genome-wide association studies (GWAS) meta-analyses of 63,415 cases of Europeans and East Asians, and identified 60 previously unreported susceptibility loci. Integrating multi-ancestry transcriptome-wide association studies (TWAS), cell type-specific TWAS, alternative splicing, and alternative polyadenylation analyses using RNA-sequencing data from normal colon tissues of 707 Europeans and 364 East Asians, we identify 441 candidate risk genes, including 324 not previously implicated by GWAS or TWAS, of which 291 were further supported by colocalization or summary-data-based Mendelian randomization analyses. These genes converge on pathways central to IBD pathogenesis (e.g., immune regulation), with over half showing transcriptional dysregulation supported by single-cell and spatial omics analyses. We further identify 44 druggable genes linked to 221 approved drugs or Phase II/III clinical-stage compounds. Our findings deepen the understanding of IBD genetics and provide insights for future research relevant to precision medicine for IBD.

## INTRODUCTION

Inflammatory Bowel Disease (IBD) comprises a group of chronic, relapsing-remitting inflammatory conditions of the gastrointestinal tract, primarily Crohn’s disease (CD) and ulcerative colitis (UC). Genome-wide association studies (GWAS) have identified more than 300 risk loci associated with IBD, primarily through large-scale studies in individuals of European and East Asian ancestry^1, 2, 3, 4, 5, 6, 7^. However, the causal genes and biological mechanisms underlying these associations remain largely unresolved. In addition, most previous GWAS have been conducted within individual ancestry groups, limiting the cross-population generalizability of genetic findings and their potential translation into broadly applicable clinical tools. Multi-ancestry analyses provide an important opportunity to distinguish shared genetic mechanisms from ancestry-enriched effects, improve the localization of association signals by leveraging differences in linkage disequilibrium (LD) structure, and enhance the relevance of genetic discoveries across diverse populations. This framework may contribute to the long-term goals of precision medicine by helping ensure that genetic discoveries and downstream biological and therapeutic insights are relevant across diverse populations, rather than being disproportionately applicable to the populations in which they were initially developed.

Transcriptome-wide association studies (TWAS) offer a powerful framework to bridge GWAS signals with gene expression, by using predictive models trained on expression quantitative trait loci (eQTLs) to identify genes whose genetically predicted expression is associated with disease risk^8, 9^. However, conventional TWAS approaches are often limited by the inclusion of non-functional variants in prediction models, reducing power and increasing false discovery rates^10, 11, 12^. To overcome these limitations, we developed sTF-TWAS, a framework that prioritizes regulatory variants within transcription factor-occupied *cis*-regulatory elements (STFCREs), significantly improving the accuracy and sensitivity of TWAS gene discovery^13, 14^. TWAS has also been extended to investigate genetically regulated post-transcriptional features such as alternative splicing (AS) and alternative polyadenylation (APA), offering additional insights into gene regulation and disease mechanisms^15^. Furthermore, single-cell RNA sequencing (scRNA-seq) enables high-resolution characterization of tissue composition and facilitates the construction of cell type-specific transcriptomic profiles^16^, thereby improving the detection of cell type-specific genetic effects^17, 18, 19, 20, 21, 22^. Integration of scRNA-seq data with computational deconvolution tools, such as CIBERSORTx^23^, has demonstrated utility in uncovering cell type-specific genetic regulation in several complex diseases^17, 18, 19, 24, 25, 26^. However, such approaches remain underutilized in IBD^27^. To date, only limited TWASs of IBD using bulk normal colon tissue has been reported^28, 29^, underscoring the need for high-resolution, multi-omics integrative analyses in this field.

To address these critical gaps, we conducted ancestry-specific and multi-ancestry GWAS meta-analyses for IBD, CD, and UC, followed by conditional analyses to distinguish novel independent signals from associations tagging established loci. We then integrated multiple complementary genomic approaches, including x-quantitative trait locus (xQTL) mapping (expression (eQTL), alternative splicing (AS-QTL), and alternative polyadenylation (APA-QTL)), TWAS, alternative splicing-wide association studies (AS-WAS), alternative polyadenylation-wide association studies (APA-WAS), cell type-specific TWAS, Fine-mapping Of CaUsal gene Sets (FOCUS) fine-mapping, Bayesian colocalization, and Summary-data-based Mendelian Randomization (SMR) analyses, to systematically prioritize candidate susceptibility genes and regulatory mechanisms. Finally, we evaluated these prioritized genes using single-cell and spatial transcriptomic datasets as complementary functional evidence and comprehensively annotated their therapeutic potential through drug-target relationships, mechanisms of action, disease-relevant pathways, and perturbational transcriptomic signatures. Together, this integrative multi-omics framework enabled the prioritization of candidate IBD susceptibility genes, including genes targeted by approved drugs or compounds currently in Phase II/III clinical trials, while providing a resource for future functional and translational studies.

## RESULTS

### Identification of genetic risk loci and target genes associated with IBD

In the meta-analysis of European populations, we included a total of 49,022 cases of IBD and 1,293,489 controls (**Supplementary Data 1**) from Veterans Affairs (VA) Million Veteran Program (MVP)^30^, the FinnGen study (R12)^31^, Pan-UK Biobank (UKB)^32^ and a GWAS meta-analysis of UK IBD Genetics Consortium (UKIBDGC) and International Inflammatory Bowel Disease Genetics Consortium (IIBDGC)^6^ (Hereafter referred as IIBDGC population). Using an inverse variance-weight fixed-effect meta-analysis (**Supplementary Methods**), we identified 16 novel independent IBD susceptibility loci that reached genome-wide significance at *P* < 5 × 10^−8^. Disease subtype-specific analyses identified an additional 9 novel loci associated with CD and 14 associated with UC (**Table 1**, **Figure 1, Supplementary Data 2, 3**).

**Figure 1.**
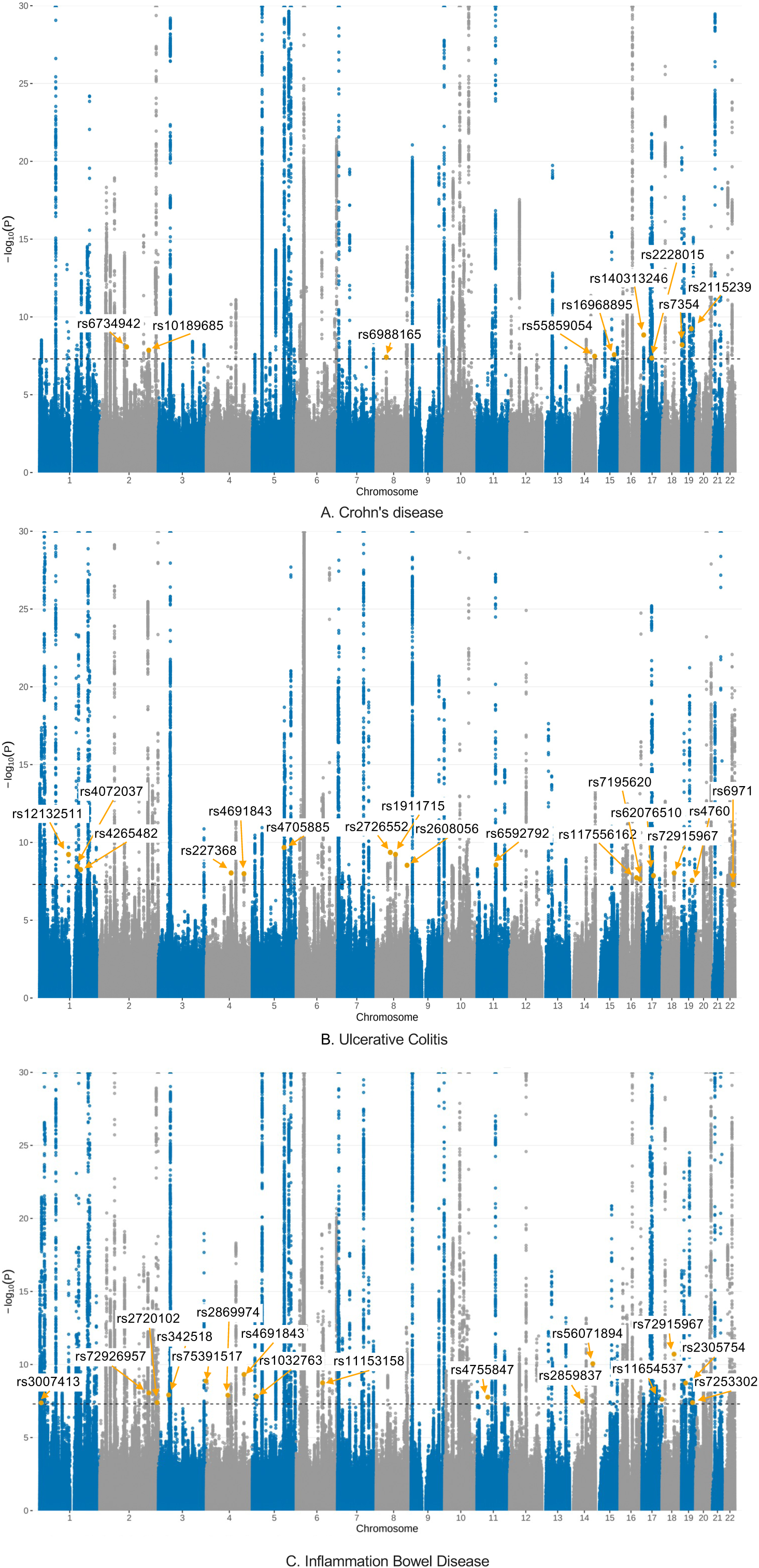
Manhattan plot of GWAS meta-analysis results in the European populations. Manhattan plots show genome-wide association signals for inflammatory bowel disease, Crohn’s disease, and ulcerative colitis, identified from GWAS meta-analysis of European ancestry (49,022 IBD cases and 1,293,489 controls). Each point represents a variant plotted by chromosomal position, with association strength shown as-log_10_(*P*). The black dashed line indicates the genome-wide significance threshold (*P* = 5 × 10^-8^). Lead variant of novel loci is highlighted and labeled. The *y*-axis was truncated to improve visualization of loci near the significance threshold; variants exceeding the upper limit are shown at the top boundary. GWAS effect estimates were combined using fixed-effect inverse-variance-weighted meta-analysis in METAL. Association *P-*values were calculated using two-sided *z*-tests, with genomic control applied to each contributing dataset and to the GWAS meta-analysis results.

**Table 1.**
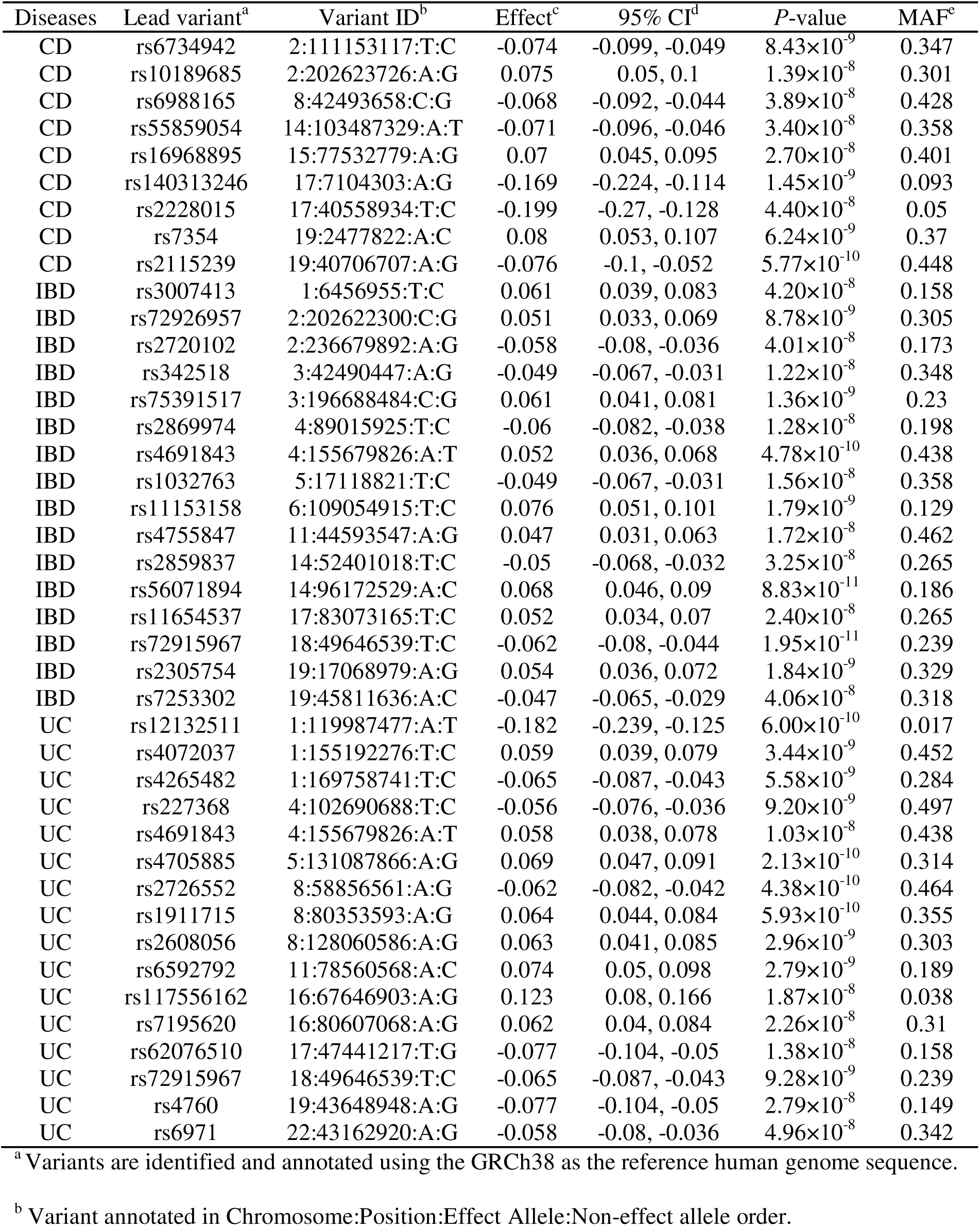
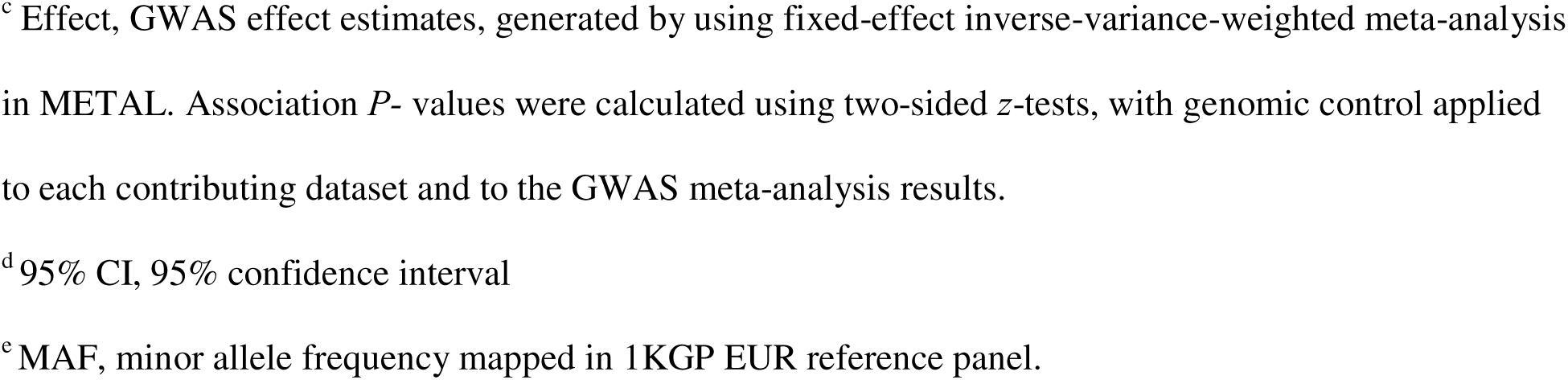
Novel genetic loci identified from GWAS meta-analysis of European ancestry.

| Diseases | Lead variant<sup>a</sup> | Variant ID<sup>b</sup> | Effect<sup>c</sup> | 95% CI<sup>d</sup> | P-value | MAF<sup>e</sup> |
| --- | --- | --- | --- | --- | --- | --- |
| CD | rs6734942 | 2:111153117:T:C | -0.074 | -0.099, -0.049 | 8.43×10<sup>-9</sup> | 0.347 |
| CD | rs10189685 | 2:202623726:A:G | 0.075 | 0.05, 0.1 | 1.39×10<sup>-8</sup> | 0.301 |
| CD | rs6988165 | 8:42493658:C:G | -0.068 | -0.092, -0.044 | 3.89×10<sup>-8</sup> | 0.428 |
| CD | rs55859054 | 14:103487329:A:T | -0.071 | -0.096, -0.046 | 3.40×10<sup>-8</sup> | 0.358 |
| CD | rs16968895 | 15:77532779:A:G | 0.07 | 0.045, 0.095 | 2.70×10<sup>-8</sup> | 0.401 |
| CD | rs140313246 | 17:7104303:A:G | -0.169 | -0.224, -0.114 | 1.45×10<sup>-9</sup> | 0.093 |
| CD | rs2228015 | 17:40558934:T:C | -0.199 | -0.27, -0.128 | 4.40×10<sup>-8</sup> | 0.05 |
| CD | rs7354 | 19:2477822:A:C | 0.08 | 0.053, 0.107 | 6.24×10<sup>-9</sup> | 0.37 |
| CD | rs2115239 | 19:40706707:A:G | -0.076 | -0.1, -0.052 | 5.77×10<sup>-10</sup> | 0.448 |
| IBD | rs3007413 | 1:6456955:T:C | 0.061 | 0.039, 0.083 | 4.20×10<sup>-8</sup> | 0.158 |
| IBD | rs72926957 | 2:202622300:C:G | 0.051 | 0.033, 0.069 | 8.78×10<sup>-9</sup> | 0.305 |
| IBD | rs2720102 | 2:236679892:A:G | -0.058 | -0.08, -0.036 | 4.01×10<sup>-8</sup> | 0.173 |
| IBD | rs342518 | 3:42490447:A:G | -0.049 | -0.067, -0.031 | 1.22×10<sup>-8</sup> | 0.348 |
| IBD | rs75391517 | 3:196688484:C:G | 0.061 | 0.041, 0.081 | 1.36×10<sup>-9</sup> | 0.23 |
| IBD | rs2869974 | 4:89015925:T:C | -0.06 | -0.082, -0.038 | 1.28×10<sup>-8</sup> | 0.198 |
| IBD | rs4691843 | 4:155679826:A:T | 0.052 | 0.036, 0.068 | 4.78×10<sup>-10</sup> | 0.438 |
| IBD | rs1032763 | 5:17118821:T:C | -0.049 | -0.067, -0.031 | 1.56×10<sup>-8</sup> | 0.358 |
| IBD | rs11153158 | 6:109054915:T:C | 0.076 | 0.051, 0.101 | 1.79×10<sup>-9</sup> | 0.129 |
| IBD | rs4755847 | 11:44593547:A:G | 0.047 | 0.031, 0.063 | 1.72×10<sup>-8</sup> | 0.462 |
| IBD | rs2859837 | 14:52401018:T:C | -0.05 | -0.068, -0.032 | 3.25×10<sup>-8</sup> | 0.265 |
| IBD | rs56071894 | 14:96172529:A:C | 0.068 | 0.046, 0.09 | 8.83×10<sup>-11</sup> | 0.186 |
| IBD | rs11654537 | 17:83073165:T:C | 0.052 | 0.034, 0.07 | 2.40×10<sup>-8</sup> | 0.265 |
| IBD | rs72915967 | 18:49646539:T:C | -0.062 | -0.08, -0.044 | 1.95×10<sup>-11</sup> | 0.239 |
| IBD | rs2305754 | 19:17068979:A:G | 0.054 | 0.036, 0.072 | 1.84×10<sup>-9</sup> | 0.329 |
| IBD | rs7253302 | 19:45811636:A:C | -0.047 | -0.065, -0.029 | 4.06×10<sup>-8</sup> | 0.318 |
| UC | rs12132511 | 1:119987477:A:T | -0.182 | -0.239, -0.125 | 6.00×10<sup>-10</sup> | 0.017 |
| UC | rs4072037 | 1:155192276:T:C | 0.059 | 0.039, 0.079 | 3.44×10<sup>-9</sup> | 0.452 |
| UC | rs4265482 | 1:169758741:T:C | -0.065 | -0.087, -0.043 | 5.58×10<sup>-9</sup> | 0.284 |
| UC | rs227368 | 4:102690688:T:C | -0.056 | -0.076, -0.036 | 9.20×10<sup>-9</sup> | 0.497 |
| UC | rs4691843 | 4:155679826:A:T | 0.058 | 0.038, 0.078 | 1.03×10<sup>-8</sup> | 0.438 |
| UC | rs4705885 | 5:131087866:A:G | 0.069 | 0.047, 0.091 | 2.13×10<sup>-10</sup> | 0.314 |
| UC | rs2726552 | 8:58856561:A:G | -0.062 | -0.082, -0.042 | 4.38×10<sup>-10</sup> | 0.464 |
| UC | rs1911715 | 8:80353593:A:G | 0.064 | 0.044, 0.084 | 5.93×10<sup>-10</sup> | 0.355 |
| UC | rs2608056 | 8:128060586:A:G | 0.063 | 0.041, 0.085 | 2.96×10<sup>-9</sup> | 0.303 |
| UC | rs6592792 | 11:78560568:A:C | 0.074 | 0.05, 0.098 | 2.79×10<sup>-9</sup> | 0.189 |
| UC | rs117556162 | 16:67646903:A:G | 0.123 | 0.08, 0.166 | 1.87×10<sup>-8</sup> | 0.038 |
| UC | rs7195620 | 16:80607068:A:G | 0.062 | 0.04, 0.084 | 2.26×10<sup>-8</sup> | 0.31 |
| UC | rs62076510 | 17:47441217:T:G | -0.077 | -0.104, -0.05 | 1.38×10<sup>-8</sup> | 0.158 |
| UC | rs72915967 | 18:49646539:T:C | -0.065 | -0.087, -0.043 | 9.28×10<sup>-9</sup> | 0.239 |
| UC | rs4760 | 19:43648948:A:G | -0.077 | -0.104, -0.05 | 2.79×10<sup>-8</sup> | 0.149 |
| UC | rs6971 | 22:43162920:A:G | -0.058 | -0.08, -0.036 | 4.96×10<sup>-8</sup> | 0.342 |

925 <sup>a</sup> Variants are identified and annotated using the GRCh38 as the reference human genome sequence.926 <sup>b</sup> Variant annotated in Chromosome:Position:Effect Allele:Non-effect allele order.
<sup>c</sup> Effect, GWAS effect estimates, generated by using fixed-effect inverse-variance-weighted meta-analysis in METAL. Association *P*- values were calculated using two-sided z-tests, with genomic control applied to each contributing dataset and to the GWAS meta-analysis results.
<sup>d</sup> 95% CI, 95% confidence interval
<sup>e</sup> MAF, minor allele frequency mapped in 1KGP EUR reference panel.

While the genomic inflation factors (λ) showed mild inflations for summary statistics (λ_CD_= 1.08, λ_UC_ = 1.10, and λ_IBD_ = 1.12), the LD score regression (LDSC) intercepts (0.97, 0.99, and 0.99 for CD, UC, and IBD, respectively) indicated that most of the observed inflation was likely attributable to polygenic signal rather than major confounding from population stratification. The quantile-quantile (Q-Q) plots of the full GWAS meta data and GWAS meta data after removing the loci that are located within 300 kilobytes (Kb) of previously identified subtype specific GWAS significant loci were presented in **Supplementary Figure 1**. The persistence of excess association signals after excluding known susceptibility loci, together with LDSC intercepts close to 1, supports the presence of widespread polygenic genetic effects while indicating that population stratification and other major confounding factors were generally well controlled. Nevertheless, given the large sample size and meta-analytic design of this study, we cannot completely exclude modest residual contributions from non-biological sources, including subtle population structure, cryptic relatedness, phenotype heterogeneity across contributing cohorts, or technical artifacts.

In the multi-ancestry meta-analysis, we aggregated a total of 63,415 IBD cases and 1,308,945 controls after combining the European data with an additional 14,393 cases of IBD and 15,456 controls from a recent GWAS of East Asian populations^7^. At genome-wide significance level, we identified additional 7 novel loci associated with IBD. Further stratified analysis revealed 7 novel genetic loci associated with CD, and 7 novel loci associated with UC (**Table 2**, **Figure 2, Supplementary Data 2, 3**). Combined genomic inflation factors (λ□=□1.07-1.10), LDSC intercepts (0.98-1.00), and the Q-Q plots (**Supplementary Figure 1**) suggests that population stratification has been generally well controlled. The Q-Q plots for individual GWAS summary statistics were presented in **Supplementary Figure 2**. Most loci showed no statistically significant heterogeneity (*P* of heterogeneity cross ancestry > 0.05) (**Supplementary Data 3**), although limited power in the East Asian component reduces the ability to exclude moderate ancestry-specific differences. The multi-ancestry Manhattan plots demonstrated that several signals were strengthened by cross-ancestry integration, while others were driven predominantly by the larger European datasets.

**Figure 2.**
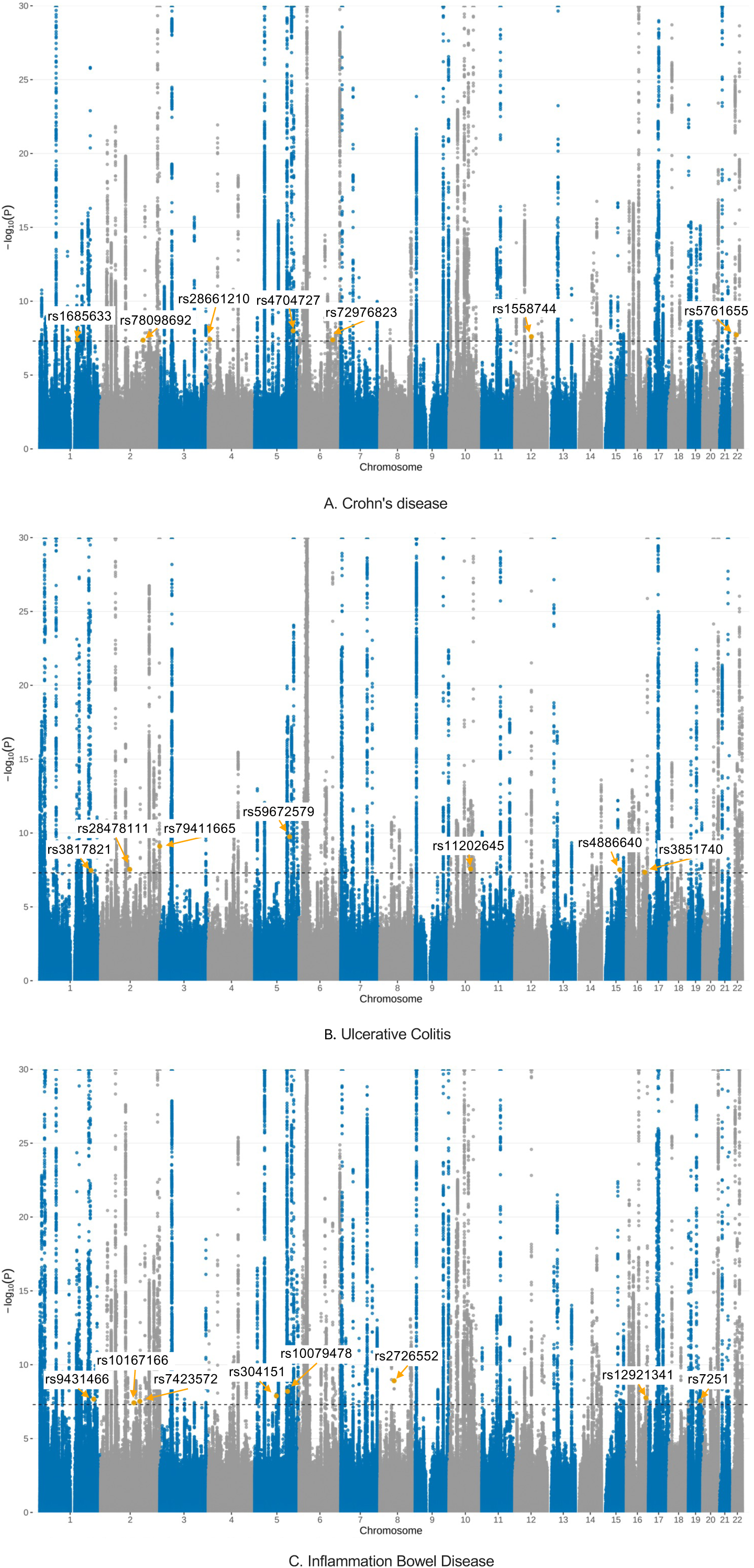
Manhattan plot of GWAS meta-analysis results across European and East Asian populations. Manhattan plots show genome-wide association signals for inflammatory bowel disease, Crohn’s disease, and ulcerative colitis, identified from GWAS meta-analysis of European and East Asian ancestry (63,415 IBD cases and 1,308,945controls). Each point represents a variant plotted by chromosomal position, with association strength shown as-log_10_(*P*). The black dashed line indicates the genome-wide significance threshold (*P* = 5 × 10^-8^). Lead variant of novel loci is highlighted and labeled. The *y*-axis was truncated to improve visualization of loci near the significance threshold; variants exceeding the upper limit are shown at the top boundary. GWAS effect estimates were combined using fixed-effect inverse-variance-weighted meta-analysis in METAL. Association *P*-values were calculated using two-sided *z*-tests, with genomic control applied to each contributing dataset and to the GWAS meta-analysis results.

**Table 2.** Novel genetic loci identified from GWAS meta-analysis of multi-ancestry.

| Diseases | Lead variant<sup>a</sup> | Variant ID<sup>b</sup> | Effect<sup>c</sup> | 95% CI<sup>d</sup> | *P*-value | MAF<sup>e</sup> |
| --- | --- | --- | --- | --- | --- | --- |
| CD | rs1685633 | 1:154319242:A:G | -0.101 | -0.136, -0.066 | 4.04×10<sup>-8</sup> | 0.106 |
| CD | rs78098692 | 2:173899186:T:C | -0.09 | -0.121, -0.059 | 4.36×10<sup>-8</sup> | 0.169 |
| CD | rs28661210 | 4:7049229:A:C | 0.063 | 0.039, 0.087 | 3.89×10<sup>-8</sup> | 0.495 |
| CD | rs4704727 | 5:156953056:T:G | 0.07 | 0.046, 0.094 | 1.01×10<sup>-8</sup> | 0.331 |
| CD | rs72976823 | 6:137698375:A:G | -0.067 | -0.091, -0.043 | 4.17×10<sup>-8</sup> | 0.288 |
| CD | rs1558744 | 12:68110812:A:G | 0.065 | 0.041, 0.089 | 2.51×10<sup>-8</sup> | 0.403 |
| CD | rs5761655 | 22:26642889:A:G | 0.066 | 0.042, 0.09 | 1.93×10<sup>-8</sup> | 0.368 |
| IBD | rs9431466 | 1:221262843:C:G | 0.047 | 0.031, 0.063 | 2.11×10<sup>-8</sup> | 0.36 |
| IBD | rs10167166 | 2:136302336:A:G | -0.065 | -0.089, -0.041 | 3.80×10<sup>-8</sup> | 0.144 |
| IBD | rs7423572 | 2:160487551:C:G | -0.053 | -0.073, -0.033 | 3.00×10<sup>-8</sup> | 0.2 |
| IBD | rs304151 | 5:88830036:T:G | 0.047 | 0.031, 0.063 | 1.30×10<sup>-8</sup> | 0.268 |
| IBD | rs10079478 | 5:134505105:T:C | -0.051 | -0.069, -0.033 | 6.37×10<sup>-9</sup> | 0.248 |
| IBD | rs2726552 | 8:58856561:A:G | -0.047 | -0.063, -0.031 | 1.23×10<sup>-9</sup> | 0.464 |
| IBD | rs12921341 | 16:80006686:T:G | -0.045 | -0.061, -0.029 | 1.73×10<sup>-8</sup> | 0.469 |
| IBD | rs7251 | 19:49659652:C:G | -0.046 | -0.062, -0.03 | 2.79×10<sup>-8</sup> | 0.333 |
| UC | rs3817821 | 1:209745158:A:T | 0.06 | 0.038, 0.082 | 3.42×10<sup>-8</sup> | 0.233 |
| UC | rs28478111 | 2:119412310:T:C | 0.065 | 0.041, 0.089 | 2.91×10<sup>-8</sup> | 0.23 |
| UC | rs79411665 | 2:241832052:T:C | 0.092 | 0.063, 0.121 | 7.91×10<sup>-10</sup> | 0.129 |
| UC | rs59672579 | 5:142866576:T:C | -0.103 | -0.134, -0.072 | 1.78×10<sup>-10</sup> | 0.099 |
| UC | rs11202645 | 10:88073835:A:G | 0.053 | 0.033, 0.073 | 2.73×10<sup>-8</sup> | 0.423 |
| UC | rs4886640 | 15:74935628:A:T | 0.064 | 0.04, 0.088 | 3.12×10<sup>-8</sup> | 0.469 |
| UC | rs3851740 | 16:75353680:T:C | 0.06 | 0.038, 0.082 | 4.55×10<sup>-8</sup> | 0.439 |

<sup>a</sup> Variants are identified and annotated using the GRCh38 as the reference human genome sequence.
<sup>b</sup> Variant annotated in Chromosome:Position:Effect Allele:Non-effect allele order.
<sup>c</sup> Effect, GWAS effect estimates, generated by using fixed-effect inverse-variance-weighted meta-analysis in METAL. Association *P*- values were calculated using two-sided z-tests, with genomic control applied to each contributing dataset and to the GWAS meta-analysis results.
<sup>d</sup> 95% CI, 95% confidence interval
<sup>e</sup> MAF, minor allele frequency mapped in 1KGP EUR reference panel.

After combining results across the European-only and multi-ancestry analyses, we identified 60 previously unreported independent loci across IBD, CD, and UC (**Table 1**, **Table 2, Supplementary Data 3**). All variants reached the genome-wide significant threshold (*P* < 5×10^-8^) were presented in **Supplementary Data 4** for European-specific meta-analysis and **Supplementary Data 5** for multi-ancestry meta-analysis.

To identify putative target genes in the 60 novel and the previously reported 317 GWAS-identified lead variants collected from literature^3, 6, 7, 30, 31, 32^ (**Supplementary Data 6; Methods**), we performed xQTL mapping, including eQTL, AS-QTL, and APA-QTL analyses, using bulk RNA-seq data from 707 European-ancestry individuals (423 from BarcUVa-Seq project^33^ and 284 from GTEx^33, 34^), and 364 Asian-ancestry individuals from our previous studies^35^ (**Methods**). In addition, we also conducted cell type-specific eQTL using cell type-specific gene expression deconvoluted from bulk colon tissue gene expression^36^ (**Methods**). In the European dataset, we tested 4,432 variant–gene expression pairs, 3,436 variant–APA pairs, 11,575 variant–AS pairs, and 2,531 cell type-specific eQTL pairs for absorptive (ABS), 987 eQTL pairs for goblet (GOB), and 2,096 eQTL pairs for stem (STM) cells, respectively. In the East Asian dataset, we tested 1,432 variant–gene expression pairs, 1,591 variant–APA pairs, 4,621 variant–AS pairs, and 694, 254, and 648 ABS-, GOB-, and STM-specific eQTL pairs, respectively.

Applying Bonferroni correction (*P* < 0.05/*n*, where n is number of tests for each analysis), we identified 245 risk genes (corresponding 295 gene-variants pairs), including 233 genes in the European population and additional 12 genes in the East Asian population (**Supplementary Data 7**). Furthermore, colocalization analysis between GWAS and xQTL signals using the R package “coloc”^37^ identified 53 candidate risk genes, supported by strong evidence of shared causal signals (posterior probability for hypothesis 4 (PP.H4) > 0.8; **Supplementary Data 8; Methods**). Of them, 13 genes are linked to lead variants from the newly identified GWAS loci.

### Prioritization of candidate risk genes through a multi-ancestry TWAS framework

We next conducted multi-ancestry TWAS, AS-WAS, and APA-WAS through ancestry-specific genetic prediction model building based on the above bulk RNA-seq data. For TWAS in European populations, 15,307 gene-level tests were performed, yielding a Bonferroni significance threshold of *P* < 3.26 × 10□□(0.05/ 15,307). Using this threshold, we identified 300 genes significantly associated with IBD risk (**Supplementary Data 9**). Meta-analysis incorporating Asian ancestry data revealed an additional 15 significant genes at a Bonferroni significance threshold of *P* < 3.04 × 10□□(0.05/ 16,447). We also identified additional genes through subtype-specific analysis. For CD, we identified 34 significant genes in the European cohort at *P* < 3.26 × 10□□ and 9 additional genes through meta-analysis at *P* < 3.04× 10□□(**Supplementary Data 9**). For UC, 35 genes were significant in the European cohort at *P* < 3. 26 × 10, with 12 additional genes identified in meta-analysis at *P* < 3.04× 10 (**Supplementary Data 9**). In total, TWAS uncovered 405 IBD-associated genes, including 376 from European ancestry and 29 additional genes identified through meta-analysis with Asian ancestry. Fine-mapping analyses using FOCUS reveal 202 candidate risk genes, including 141 with posterior inclusion probability (PIP) > 0.5 from FOCUS (**Supplementary Data 10**) and an additional 61 from regions beyond FOCUS analyses (**Supplementary Data 11**). Here, PIP > 0.5 represented a prioritization criterion and not as definitive evidence of causality.

For APA-WAS, we identified 67 genes associated with IBD, including 61 in European populations at *P* < 1.14 × 10□^5^ (0.05/4,380) and additional 6 genes at *P* < 8.67 × 10□□(0.05/5,763) from meta-analysis of European and East Asian populations (**Supplementary Data 12**). For CD, we identified 10 additional significant genes associated with CD in the European cohort at *P* < 1.14 × 10□^5^, while no significant genes were detected from the meta-analysis (**Supplementary Data 12**). For UC, APA-WAS identified 6 additional significant genes in European ancestry at *P* < 1.14 × 10□^5^ and 4 additional genes from meta-analysis at *P* < 8.67 × 10□□(**Supplementary Data 12**). In total, APA-WAS identified 87 genes, including 77 from European ancestry and 10 additional genes through meta-analysis (**Supplementary Data 12**). Fine-mapping analysis prioritized 56 prioritized candidate risk genes, including 43 with PIP > 0.5 from FOCUS analyses (**Supplementary Data 13**) and 13 additional genes with the smallest APA-WAS *P*-values within ±1 Megabyte (Mb) loci (**Supplementary Data 14**).

For AS-WAS, we identified 221 genes associated with IBD, including 207 in European populations at *P* < 2.39 × 10□□(0.05/20,914) and additional 14 genes from meta-analysis of European and East Asian populations at *P* < 2.14 × 10□□(0.05/23,402) (**Supplementary Data 15**). For CD, 24 genes were significant in European ancestry at *P* < 2.39 × 10□□, and 8 more were identified via meta-analysis at *P* < 2.14 × 10□□(**Supplementary Data 15**). For UC, 23 genes were significant in European ancestry at *P* < 2.39 × 10□□, and an additional 5 genes were discovered through meta-analysis at *P* < 2.14 × 10□□(**Supplementary Data 15**). Altogether, AS-WAS identified 281 genes, comprising 257 from European ancestry and 24 additional genes from meta-analysis with Asian ancestry. Fine-mapping underscored 122 prioritized candidate risk genes, including 98 with PIP > 0.5 from FOCUS analyses (**Supplementary Data 16**) and 24 additional genes with the smallest AS-WAS *P*-values within ±1 Mb loci (**Supplementary Data 17**).

For cell type-specific TWAS, we first estimated the heritability contributed by Single Nucleotide Polymorphisms (SNPs) for three major cell type, ABS cells, GOB, and STM using stratified LDSC (sLDSC) analysis^38^ (**Supplementary Methods**). The analysis revealed substantial heritability contributions from these cell types (**Supplementary Data 18**). For CD as an example, among the European ancestry, heritability was contributed to ABS cells with 65.19% (standard error (SE): 0.06), GOB with 49.36% (SE: 0.05), and STM with 21.95% (SE: 0.03).

We then performed cell-type specific TWAS using prediction models built for three major cell types (ABS, GOB, and STM). Statistical significance was defined using cell type-, ancestry-, and analysis-specific Bonferroni thresholds, which were shared across IBD, CD, and UC within each corresponding analysis. In European ancestry, the Bonferroni significance thresholds were *P* < 8.02 × 10□□for ABS cells (0.05/6,230), *P* < 6.47 × 10□^5^ for GOB cells (0.05/772), and *P* < 8.89 × 10□^6^ for STM cells (0.05/5,619). For the multi-ancestry meta-analysis, the Bonferroni significance thresholds were *P* < 7.27 × 10□□for ABS cells (0.05/6,872), *P* < 5.16 × 10□^5^ for GOB cells (0.05/969), and *P* < 7.97 × 10□^6^ (0.05/6,276) for STM cells. Using these thresholds, we identified 220 genes significantly associated with IBD (**Supplementary Data 19**), with 206 from in European and an additional 14 genes through meta-analysis with Asian ancestry (**Supplementary Data 19**). For the subtype CD, we identified 34 significant genes in the European cohort (**Supplementary Data 19**) and 3 additional genes via meta-analysis. For the subtype UC, we observed 31 additional significant genes in the European cohort and 8 more genes through meta-analysis. In total, the cell type-specific TWAS analysis uncovered 296 genes associated with IBD, CD and UC, including 273 from European ancestry and 23 additional genes identified through cross-ancestry meta-analysis (**Supplementary Data 19**). Fine-mapping underscored 211 prioritized candidate risk genes, including 156 with PIP > 0.5 from FOCUS analyses (**Supplementary Data 20**) and 55 additional genes with the smallest cell type-specific TWAS *P*-values within 1 Mb loci (**Supplementary Data 21**).

After pooling results across the above analyses, we uncovered 441 prioritized candidate risk genes that are associated with IBD, CD and UC. Of these, 315 genes have not been reported in previous TWAS^28, 29^, with 248 from TWAS, APA-WAS, and AS-WAS (**Figure 3**) and 134 from cell type-specific TWAS (**Supplementary Figure 3**) with 67 genes shared by bulk tissue and single cell studies. Among these, we identified 88 genes located in 30 GWAS loci that had not been previously reported (**Supplementary Data 22**). After combining the results with xQTL mapping in GWAS risk loci, we identified 441 prioritized candidate risk genes for IBD. Of them, 324 genes have not been reported in previous TWAS^28, 29^ and GWAS^3, 6, 7^ (**Supplementary Data 23**). Of the 441 prioritized candidate risk genes, we found that analyses from these molecular traits largely provide complementary contributions to the discovery of risk genes (**Supplementary Figure 4**). After adjusting for the number of tests in the corresponding ancestry, panel, and diseases, 283 prioritized candidate risk genes were further supported by SMR analysis (**Supplementary Data 24**). Colocalization analysis identified 8 additional genes not detected by SMR analysis (**Supplementary Data 25**). Taking together, 291 (65.99%) prioritized candidate risk genes were supported by either SMR or colocalization analysis (PP.H4 > 0.8) in European and East Asian populations.

**Figure 3.**
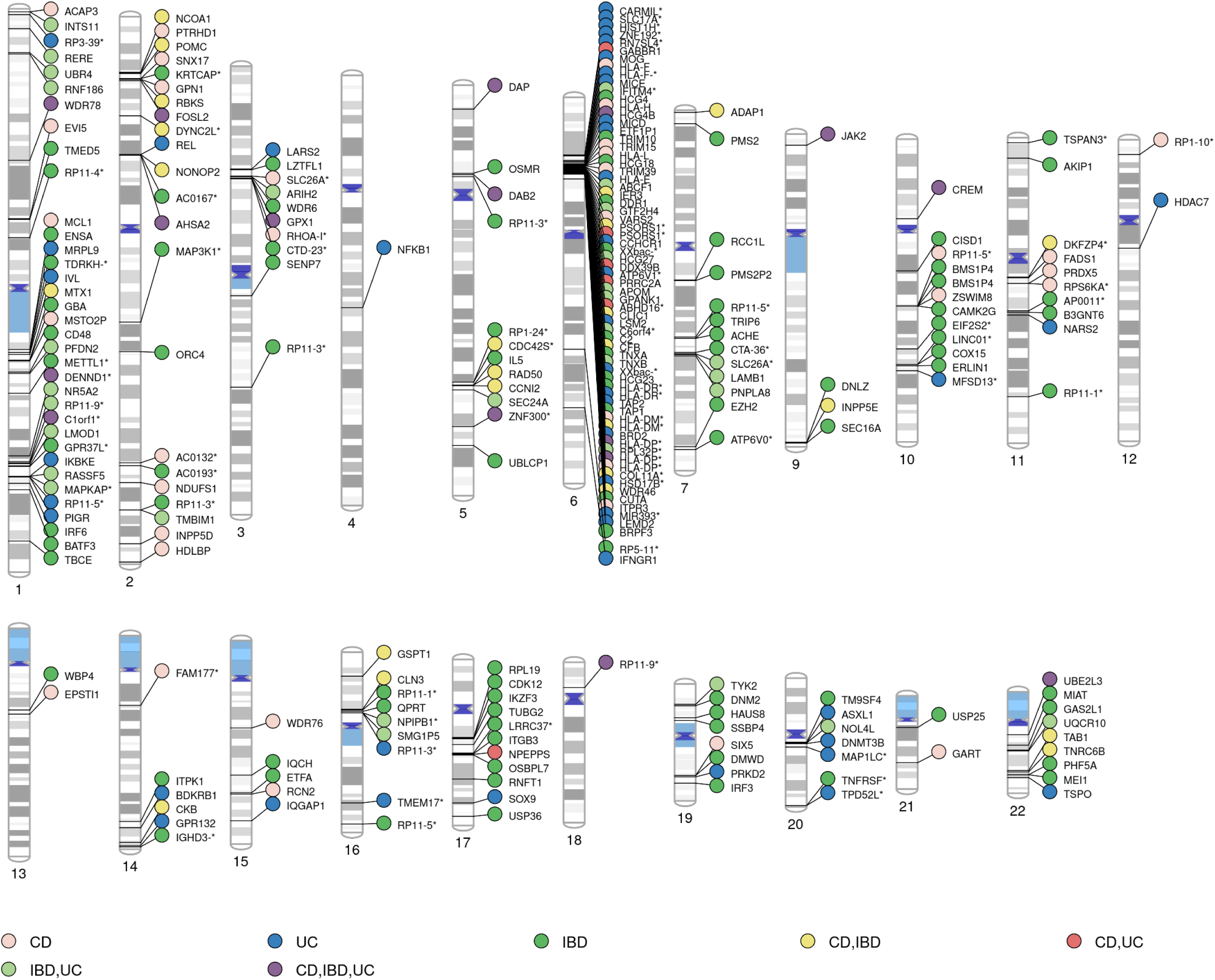
Novel prioritized candidate risk genes identified across IBD, CD, and UC from TWAS, APA-WAS, and AS-WAS analyses. 248 novel prioritized candidate risk genes identified by transcriptome-wide association studies (TWAS), alternative polyadenylation-wide association studies (APA-WAS) and alternative splicing-wide association studies (AS-WAS) are displayed according to their chromosomal positions. Genes are grouped according to their phenotype associations: “IBD” (unique to inflammatory bowel disease), “CD” (unique to Crohn’s disease), “UC” (unique to ulcerative colitis), “CD,IBD” (shared by CD and IBD), “CD,UC” (shared by CD and UC), “IBD,UC” (shared by IBD and UC), and “CD,IBD,UC” (shared by all three phenotypes). Gene names are truncated with remaining characters replaced by an asterisk (*) due to space constraints. Associations were evaluated using two-sided Wald *z*-tests implemented in sTF-TWAS. Statistical significance was determined separately for each analysis using a Bonferroni-corrected threshold of *P*/*n* where *n* was the number of models tested in the corresponding disease-and ancestry-specific analysis.

### Functional characterization of prioritized candidate risk genes

Functional enrichment analyses on the 441 prioritized candidate risk genes revealed significant enrichment in immune and inflammatory pathways that are strongly implicated in IBD pathogenesis and colorectal carcinogenesis. The most prominent pathways included cytokine signaling, interferon signaling (type I and type II), *IL-6/JAK/STAT3* signaling, *IL-2/STAT5* signaling, *Th17* cell differentiation, T-cell receptor (*TCR*) signaling, *PD-1* signaling, antigen processing and presentation, and the intestinal immune network for *IgA* production (**Supplementary Data 26; Supplementary Methods**).

These enriched pathways recapitulate major biological mechanisms previously implicated by IBD genetics, including Microbial Sensing & Innate Immunity, Autophagy, *IL-23/Th17* Signaling, *NF-*κ*B* Signaling, Epithelial Barrier, Antigen Presentation (*HLA/ERAP*), T Cell Regulation, and Leukocyte Trafficking **(Supplementary Data 27**). These include key regulators of innate immune sensing and autophagy such as *ATG16L1*; *Th17*/*IL-23* signaling genes including *IL23R*, *STAT3*, *JAK2*, and *TYK2*; NF-κB signaling components such as *NFKB1* and *REL*; epithelial barrier and mucosal integrity genes like *MUC1* and *GNA12*; and antigen processing and presentation molecules within the *HLA* region (e.g., *HLA-DQA1, HLA-DRB1, TAP1*, *ERAP2*). In addition, beyond confirming established IBD biology, our integrative analyses further prioritized additional candidate pathways and susceptibility genes that may refine the molecular mechanisms underlying IBD within related immune and epithelial networks. These included pathways not represented in our curated set of established IBD mechanisms, such as *HIF-1* signaling, complement activation, apoptosis, and intestinal *IgA*-mediated mucosal immunity. We also prioritized candidate genes involved in interferon-γ response (*PSMB9*, *SELP*), antigen cross-presentation (*SEC24A*), *HIF-1* signaling (*PRKCB*, *IFNGR1*, *AKT2*), and cell adhesion and co-stimulatory immune signaling (*CD40*). Rather than identifying entirely new classes of biological pathways, these findings extend current understanding of IBD by prioritizing specific susceptibility genes, regulatory mechanisms, and molecular processes supported by convergent evidence from GWAS, TWAS, AS, APA, and cell type-specific analyses.

### Prioritized candidate risk genes supported by scRNA-seq and spatial transcriptomics data

We next analyzed scRNA-seq datasets on normal colon tissues from 18 non-IBD controls and inflamed colon tissues from 65 CD patients^27^ (**Supplementary Methods**) using differential expression analysis on three major cell types (ABS, GOB, and STM). We provided evidence that the prioritized candidate risk genes (*P* = 5.98 × 10^-13^), or genes stratified by cell type-specific TWAS (*P* = 8.99 × 10^-5^), bulk-level TWAS approaches (TWAS, APA-WAS, and AS-WAS (*P* = 2.82 × 10^-16^), and were significantly enriched among the differentially expressed genes (DEGs) between non-IBD controls and CD patients (**Supplementary Data 28; Supplementary Methods**). For example, the genes identified through ABS-specific TWAS were enriched among DEGs in ABS cells (*P* = 3.97 × 10^-4^) (**Supplementary Data 28**). Additionally, risk genes from bulk-level TWAS approaches were enriched among DEGs in all three cell types: ABS (*P* = 1.01 × 10^-17^), GOB (*P* = 2.35 × 10^-7^), and STM (*P* = 1.06 × 10^-17^) (**Supplementary Data 28**).

Among the 441 prioritized candidate risk genes, 214 genes (48.53%) were supported by differential expression analysis (false discovery rate < 0.05, with consistent association direction relative to TWAS or xQTL results, providing additional evidence of transcriptional dysregulation involved in IBD pathogenesis (**Supplementary Data 29 – 31**). Specifically, 79 genes (17.91%) were identified through single-cell differential expression analysis by comparing the expression between 18 non-IBD controls and 65 CD patients (**Supplementary Data 29**), while 189 genes were supported by cell type-specific expression from spatial transcriptomics data, including 135 (30.61%) genes not detected in the single-cell differential expression analysis (**Supplementary Data 30**, **31**).

### Characterizing drug-target relationships and clinically advanced compounds

Using data from DrugBank^39^, ChEMBL^40^, and the TTD^41^, we comprehensively annotated drug-target relationships involving the prioritized candidate genes and approved or clinical-stage compounds. Among 441 genes, we identified 57 genes encoding druggable proteins, potentially targeted by 758 approved drugs or drugs in any stages of clinical trials (**Supplementary Data 32**). Mechanism-of-action annotations were available for 143 drug-target interactions. Most of which were associated with reduced target activity, including 14 agonists and 3 activators. These annotations provide additional information on interpreting therapeutic directionality and highlighting drug-target pairs for further investigation. To prioritize candidates with greater clinical relevance, we filtered the initial set of 57 genes and 758 drugs to retain drugs that were approved or had reached phase II or III clinical trials. This resulted in 44 genes linked to 221 drugs (**Figure 4**), providing a set of clinically advanced drug-target pairs for further mechanistic and translational evaluation in CD, IBD, and UC. Of note, our results identified Sulfasalazine, a commonly used treatment for IBD that is annotated as inhibiting *NFKB1*, and Tenapanor, a treatment for irritable bowel syndrome with constipation that is annotated as inhibiting *SLC9A3*. These findings nominate potentially relevant drug–target relationships for further investigation. Among the druggable genes, 15.8% were associated with IBD, CD, or UC in DisGeNET^42^. Pathway enrichment analysis demonstrated that the druggable genes were preferentially enriched in therapeutically actionable pathways, including immune signaling pathways (cytokine, chemokine, T-cell receptor, Toll-like receptor, interleukin, and *PD-1* checkpoint signaling), inflammatory signaling cascades (*NF-*κ*B*, *JAK–STAT*, *PI3K– AKT*, *MAPK*/*RAF*/*RAS*, *VEGF*, *Notch*, and *Wnt/*β*-catenin* signaling), and extracellular matrix remodeling pathways (focal adhesion and extracellular matrix organization) (**Supplementary Data 33**).

**Figure 4.**
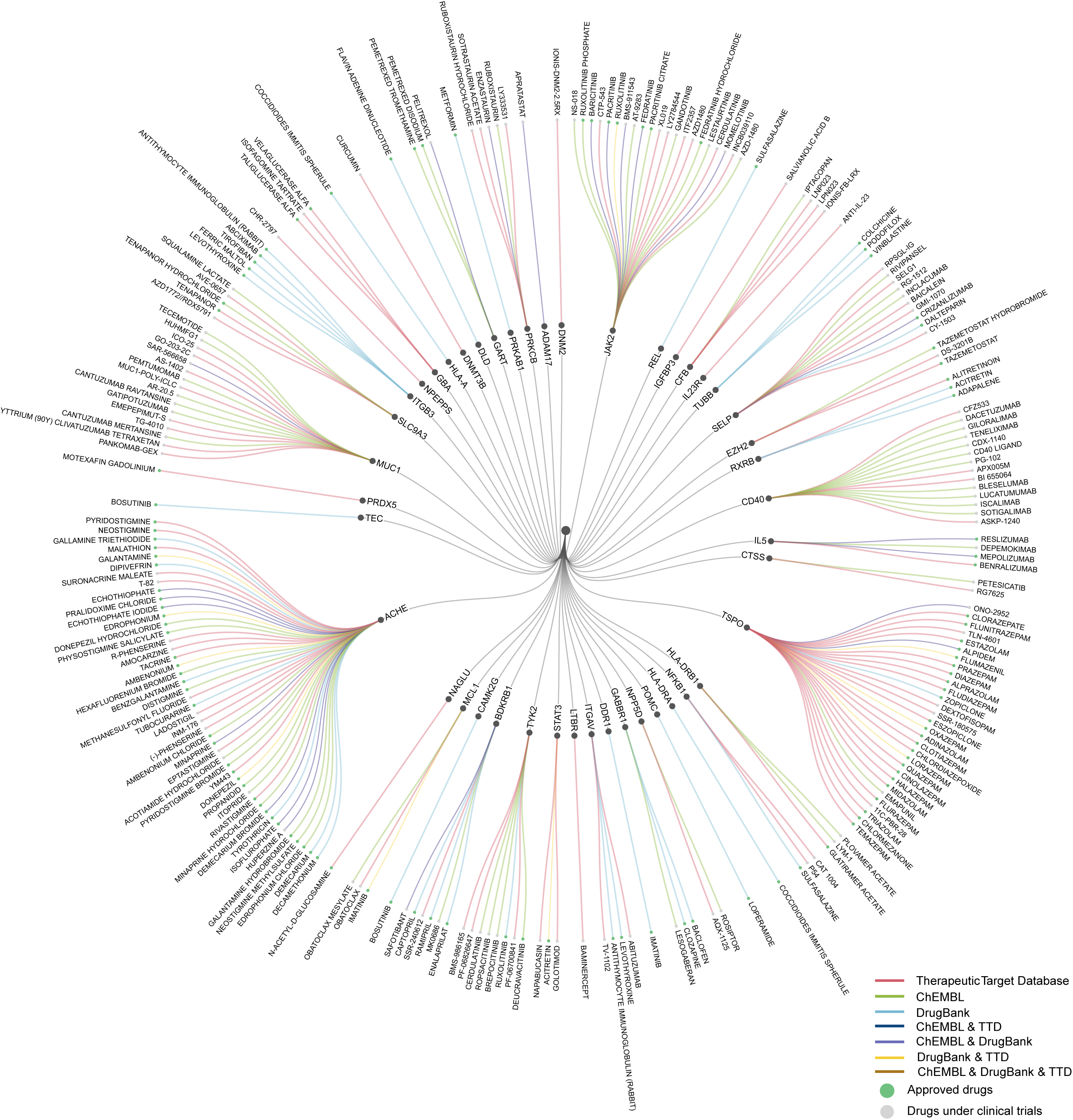
Circular plot of potential druggable genes and associated drugs for IBD treatment. Circular representation of 44 prioritized candidate risk genes encoding potentially druggable proteins and the 221 associated compounds that were either approved or had reached phase II or III clinical trials. Drug-target relationships were annotated using DrugBank³, ChEMBL³ and the Therapeutic Target Database (TTD). From the innermost to the outermost layer, the plot displays the associated phenotype, Crohn’s disease (CD), inflammatory bowel disease (IBD) or ulcerative colitis (UC), the prioritized druggable proteins and their associated compounds. Connecting lines indicate annotated drug– target relationships. The intermediate ring lists the prioritized protein targets, and the outer ring lists the associated drugs. Grey lines connect the disease group to prioritized targets, whereas colored lines connect targets to drugs and indicate the database or combination of databases supporting each drug-target relationship: red, TTD; light green, ChEMBL; light blue, DrugBank; dark blue, ChEMBL and TTD; purple, ChEMBL and DrugBank; yellow, DrugBank and TTD; and orange, ChEMBL, DrugBank and TTD. Green drug nodes indicate approved drugs, and grey drug nodes indicate drugs undergoing clinical trials. The network presents annotated drug-target relationships and does not indicate demonstrated efficacy for treating IBD, CD or UC.

To further explore drug-repositioning opportunities, we compared TWAS-inferred disease-associated transcriptional profiles with chemical perturbation signatures from large intestine-derived cell lines in the iLINCS^43^ database (**Supplementary Methods**). Seven drugs showed significant negative correlations with IBD-associated transcriptional profiles in large intestine–derived cell lines (Pearson correlation, *P* < 0.05; **Supplementary Data 34**): Baricitinib, Fedratinib, and Ruxolitinib, targeting *JAK2*; Donepezil, Neostigmine, and Pyridostigmine, targeting *ACHE*; and Bosutinib, targeting *CAMK2G*. These results suggest that these compounds may partially reverse genetically inferred disease-associated transcriptional programs.

## DISCUSSION

In this study, we identified 60 previously unreported loci associated with IBD through a meta-analysis of multi-ancestry GWAS data, thereby significantly expanding the known genetic architecture of IBD. To prioritize candidate risk genes, we integrated large-scale, multi-ancestry, and multi-omics datasets using an innovative sTF-TWAS approach that incorporated both bulk and single-cell expression data. Fine-mapping and colocalization analyses further supported the identification of 441 prioritized candidate IBD risk genes, including 324 not previously reported genes. Among these, several genes (e.g., *NFKB1*^44^ and *STAT3*^45, 46^) have previously been implicated as potential risk genes due to their proximity to GWAS risk loci. Our TWAS and eQTL analyses provide convergent statistical evidence supporting the prioritization of these genes as candidate risk genes. Our findings significantly increase the number of putative susceptibility genes and deepen our understanding of the cellular mechanisms driving IBD pathogenesis.

The different xQTL modalities provided complementary insights into the regulatory mechanisms underlying IBD susceptibility. Bulk eQTLs primarily implicated changes in overall transcript abundance, whereas AS-QTLs and APA-QTLs highlighted post-transcriptional regulatory mechanisms. Epithelial cell type-specific eQTLs further localized regulatory effects to ABS, GOB, and STM-cell populations within the colonic epithelium. Rather than considering any individual xQTL modality as intrinsically more informative, we prioritized risk genes for potential evidence of causality, supported by convergent evidence across multiple orthogonal approaches, including TWAS, xQTL mapping, Bayesian colocalization, and SMR analyses.

Although the vast majority of associations from our omics analyses were directionally consistent across populations and expression reference panels, a small proportion exhibited opposite effect directions. Such inconsistencies may arise from differences in LD structure, allele frequencies, ancestry-specific xQTL architecture, expression prediction models, or statistical uncertainty, rather than reflecting true biological discordance. Therefore, genes with discordant directions should be interpreted cautiously and require independent replication. More broadly, because our findings are based largely on *in silico* analyses, functional validation is required regardless of the direction or consistency of the observed associations to clarify the underlying regulatory mechanisms, including potential ancestry-specific effects.

Although our study primarily focused on regulatory mechanisms in normal colon tissue and epithelial cell populations, we additionally explored immune cell-specific *cis*-eQTLs using the OneK1K^25^ single-cell resource as an orthogonal source of functional annotation. Lead GWAS variants showed regulatory associations with genes across multiple immune cell types, suggesting that a subset of IBD susceptibility variants may exert their effects through immune-mediated pathways in addition to epithelial mechanisms (**Supplementary Data 35, 36**). These observations complement our colon tissue analyses and further support the biological plausibility of several prioritized genes. However, because the immune-cell analyses were intended as supportive annotation rather than systematic discovery and were evaluated using nominal significance thresholds, they should be interpreted cautiously and warrant validation in larger disease-relevant immune-cell datasets.

Beyond genes mapped to canonical immune and epithelial barrier pathways, we identified a substantial number of novel risk genes potentially involved in underexplored disease mechanisms. For example, genes involved in vesicular protein trafficking within the early secretory pathway (e.g., *TMED5*)^47^ is important in maintaining Golgi apparatus structure and facilitating the transport of glycosylphosphatidylinositol-anchored proteins. *TMED* protein family has been suggested in regulating innate immune system signaling pathways, cell differentiation, and secretory route^47^. Disruption of *TMED* proteins may contribute to immune disease by disrupting cellular secretory processes. Genes involved in cell adhesion and signal transduction (e.g., *ITGB3*)^48^ suggest an important role in reactive oxygen species-induced migration and invasion in colorectal cancer. *ITGB3* was also d to enrich in intestinal epithelial cells, whose elevation can induce cell senescence signatures in colonic epithelial cells^49^. It has been suggested to potentially associated with the development of IBD^50^. Considering the role of oxidative stress and epithelial turnover in chronic intestinal inflammation, dysregulation of *ITGB3* may contribute to persistent mucosal damage in IBD patients. Additional candidates, such as *NUCKS1*, essential in genome stability and metabolic regulation, are involved in DNA damage response and metabolism, inflammatory immune response, and microRNA^51^. These characteristics suggest that it may modulate transcriptional and post-transcriptional programs relevant to sustained inflammation in IBD. Experimental study has indicated that *NUCKS1* may be involved in DNA double-strand breaks repair and/or inflammatory signaling in UC^52^. In addition, *GSDMB* was found to be a risk gene for IBD from differential splicing analysis. *GSDMB* was found to be involved in chronic mucosal inflammatory disorders, and SNPs within the protein coding and transcriptional regulatory regions of *GSDMB* are associated with increased susceptibility of acquiring immune-mediated disorders, such as IBD^2, 53^. Our discovery revealed additional key biological processes relevant to IBD, including epithelial–immune crosstalk, stress responses, and metabolic reprogramming, providing new opportunities for functional validation and therapeutic development.

To explore the translational potential of our findings, we integrated drug-target interaction databases with the prioritized susceptibility genes. This analysis identified 44 candidate genes targeted by 221 approved drugs or compounds in Phase II/III clinical trials, providing a systematic resource for therapeutic target prioritization. Several identified drugs, including furosemide (targeting *GPR35*) and metformin (modulating *PRKAB1*-mediated *AMPK* signaling), are commonly prescribed for comorbid conditions in individuals with IBD. Among the compounds prioritized by iLINCS perturbational analyses, baricitinib and ruxolitinib, both targeting *JAK2*, demonstrated strong mechanistic plausibility given the established role of *JAK–STAT* signaling in IBD pathogenesis. However, these findings should be considered hypothesis-generating rather than evidence of therapeutic efficacy. Future validation using experimental models, proteomic analyses, real-world data, and prospective clinical studies will be necessary to determine their therapeutic value.

Several limitations should be acknowledged. Our GWAS meta-analyses were based on summary-level statistics from multiple cohorts. Although rigorous quality control, near-unity LDSC intercepts, and improved Q-Q plots after excluding known loci suggest that the modest genomic inflation primarily reflects the polygenic architecture of IBD rather than substantial confounding, we cannot completely exclude residual effects from subtle population structure, phenotype heterogeneity, limited sample overlap, or technical artifacts. Although phenotype definitions varied modestly across cohorts because of differences in healthcare systems and ascertainment strategies, all participating studies employed validated clinical-or registry-based diagnostic criteria for IBD, CD, and UC. Differences in phenotype ascertainment may contribute to residual heterogeneity. While non-differential phenotype misclassification would generally be expected to attenuate association signals, its extent and direction cannot be fully determined from summary-level data. Although our integrative framework combined TWAS, xQTL analyses, Bayesian colocalization, SMR, FOCUS fine-mapping, immune cell-specific eQTLs, and single-cell and spatial transcriptomic data to prioritize candidate genes, these complementary approaches provide convergent statistical evidence rather than definitive proof of causality. Furthermore, because the East Asian GWAS sample size remains substantially smaller than the European cohort, FOCUS was performed separately within each ancestry. Also, the use of ancestry-matched 1000 Genomes Project (1KGP) reference panels may not fully capture LD patterns across all contributing cohorts. Larger multi-ancestry datasets and improved LD reference panels will further refine causal gene prioritization. Also, statistical prioritization in *HLA* region may reflect signals shared across multiple correlated genes and variants rather than an independent effect attributable to a single gene. Although ancestry-matched LD references were used, residual LD complexity may still affect the results. We therefore report *HLA* associations separately and do not regard them as definitive evidence of single-gene causality. Regarding the gene set enrichment analysis, the observed enrichment of genetically prioritized genes among differentially expressed genes in CD colon biopsies indicates that these genes are transcriptionally perturbed in disease-relevant tissue. However, this overlap should not be interpreted as evidence of CD-specific or etiologic effects, as the expression changes may reflect general inflammation, tissue remodeling, or altered cellular composition. Comparisons with non-IBD inflammatory colitis controls and non-inflamed IBD tissue will be required to determine whether these transcriptional alterations are specific to CD. Finally, our therapeutic prioritization was based primarily on genetically predicted gene expression rather than direct protein measurements; therefore, the proposed drug targets require additional proteomic, functional, and clinical validation before translation into clinical practice.

## METHODS

This study involved secondary analyses of previously collected, de-identified or publicly available GWAS summary statistics and transcriptomic, genetic, single-cell and spatial transcriptomic datasets. No new participants were recruited, no new biospecimens were collected, and no identifiable individual-level information was accessed for the present analyses. Accordingly, no additional ethics approval or informed consent was required for this secondary analysis; ethical oversight and consent for the original data collection were governed by the respective source studies and repositories.

### GWAS cohorts in both European and Asian populations

In the meta-analysis of European ancestry, we obtained the GWAS summary statistics from three large biobanks: VA MVP^30^, the FinnGen study (R12, November 2024)^31^, Pan-UKB^32^, and a published GWAS meta-analysis of IBD^6^ from IIBDGC population. VA MVP is one of the largest US national cohorts containing comprehensive genotype and phenotype data from the VA electronic health record (EHR), with GWAS summary statistics released in July 2024. IBD cases from VA EHR were defined by phecodes curated by the MVP Data Core. For each phecode, participants with ≥2 phecode-mapped International Classification of Diseases (ICD)-9 or ICD-10 codes were defined as cases. Controls were defined as those with no instance of a phecode-mapped ICD-9 or ICD-10 code. The current meta-analysis included 7,837 individuals with IBD (N_control_ = 438,916), 3,402 individuals with CD (N_control_ = 446,631), and 5,229 individuals with UC (N_control_ = 442,102) from MVP GWAS summary statistics. FinnGen is a large public-private collaboration project within Finland and international partners. It combines and analyzes large-scale genomic and health record data from Finnish biobank and national health registries. We included the GWAS summary statistics from FinnGen in our analysis, with a total of 10,960 cases of IBD (N_control_ = 489,388), 2,489 cases of CD (N_control_ = 497,622), and 7,220 cases of UC (N_control_ = 492,160). In FinnGen, it defines cases by requiring an official Kela medication reimbursement code combined with longitudinal diagnostic validation across ICD-8, ICD-9, and ICD-10 registry histories. It also requires at least two hospital registry events for the specific CD and UC sub-phenotypes^31^. In addition, we obtained the summary statistics of IBD from the Pan-UKB project, with a total of 5,183 cases of IBD, 2,008 cases of CD, 3,670 cases of UC, and 330,270 controls. For the Pan-UKB project, cases are primarily classified by phecodes. Finally, we included a summary statistic from a GWAS meta-analysis of IIBDGC population, which contained a total of 25,042 cases of IBD (N_control_ = 34,915), 12,194 cases of CD (N_control_ = 28,072), and 12,366 cases of UC (N_control_ = 33,609). The diagnosis of IBD was followed by endoscopic, histopathological, and radiological criteria. In addition to the above European individuals, we included additional East Asian populations^7^, including 14,393 cases of IBD, 7,372 cases of CD, 6,862 cases of UC, and 15,456 controls (**Supplementary Data 1**) in our multi-ancestry GWAS meta-analysis. IBD cases were diagnosed by either the European Evidence-based Consensus or the European Crohn’s and Colitis Organisation and the European Society of Gastrointestinal and Abdominal Radiology (ECCO-ESGAR) Guideline for Diagnostic Assessment in IBD in the East Asian cohorts^7^.

Clinical characteristics, radiological and endoscopic screenings, and histological features were employed in diagnosis^7^. To consider phenotype comparability, we conducted a meta-analysis of GWAS summary statistics based on disease definitions (IBD, CD, and UC) across cohorts and did not combine different gastrointestinal phenotypes. Additional details regarding cohort-specific quality control, phenotype definitions, association analysis, and genomic inflation assessment are provided in the **Supplementary Methods.**

### Novel locus definition

Novelty was evaluated separately for IBD, CD, and UC. Lead variants located within 500 Kb of a previously reported susceptibility locus for the corresponding phenotype were considered part of the known locus and were not classified as novel. Candidate lead variants located 500 Kb to 1 Mb from a previously reported lead variant were evaluated by conditional analysis^54^ using the corresponding ancestry-and phenotype-specific GWAS meta-analysis summary statistics and ancestry-matched 1KGP^55^ for LD estimation. A locus was retained as novel only if its lead variant was located more than 1 Mb from any previously reported locus for the corresponding phenotype or remained independently associated after conditioning on the nearest reported lead variant (*P*_conditional_ < 5 × 10). Loci previously reported for one IBD subtype but independently associated with another subtype in the present study were classified as novel subtype-associated loci. A detailed description of the novel locus definition, ancestry-specific LD clumping strategy, and conditional analysis, are provided in the **Supplementary Methods**.

### Identifying putative target genes for GWAS risk loci through xQTL and colocalization analyses

To identify putative target genes associated with both novel and previously established GWAS-identified risk variants, we performed xQTL mapping using normalized bulk gene expression, AS, APA, and cell type-specific gene expression data. Bulk gene expression data were obtained from 707 European-ancestry samples, including 423 from the BarcUVa-Seq project^33^ and 284 from the Genotype-Tissue Expression (GTEx) project^33, 34^, along with 364 Asian-ancestry samples from our previous study^35^. In addition to gene expression, we generated molecular phenotypes for AS and APA from the same RNA-seq datasets. Cell type-specific gene expression profiles were obtained from 33,806 epithelial cells from normal colon tissue using the Colon Molecular Atlas Project (COLON MAP)^56^. These bulk RNA-seq datasets were used for xQTL mapping and for constructing ancestry-specific TWAS prediction models.

Details regarding genotyping and RNA-seq data processing, normalization, covariate adjustment, and model construction were provided in the **Supplementary Methods**. Linear regression analyses were conducted between each risk variant and molecular traits (eQTL, AS-QTL, APA-QTL) or cell type-specific eQTL) for genes located within ±1 Mb of the variant, adjusting for potential confounders as described in the preceding section. For cell type-specific eQTL analyses, we focused on expression data from ABS, GOB, and STM cells. Since the majority of GWAS variants were identified in European populations, the corresponding xQTL analyses were primarily conducted using the above data from individuals of European ancestry. For additional risk variants identified through meta-analysis of European and East Asian populations, we also performed xQTL analyses using the above data from the above East Asian individuals.

Statistical significance for xQTL associations was determined using a Bonferroni-corrected threshold of *P* < 0.05/*n*, where *n* denotes the number of association tests performed within each disease-, ancestry-, and xQTL-specific analysis. For these significant xQTLs, we conducted colocalization analyses to evaluate the likelihood of shared causal variants between each molecular trait and the GWAS signal. We extracted GWAS summary statistics and the corresponding xQTL data for regions spanning ±500 Kb around each GWAS lead variant, capturing variants in LD with the lead signal. Analyses were restricted to regions containing at least 50 overlapping variants, consistent with recommendations for colocalization analyses. We employed default priors and utilized the ‘coloc.abf’ function to calculate the posterior probability of colocalization. PP.H4, which reflects a shared causal variant between the two traits, was used to assess colocalization. A PP.H4 > 0.8 was considered evidence of colocalization. ABF colocalization was conducted using the package “coloc” in R^37^.

### TWAS, AS-WAS, APA-WAS and cell type-specific TWAS

#### Genetic prediction models

We applied our sTF-TWAS framework^13, 14^ to train prediction models for gene expression, AS, and APA levels. This approach utilizes *cis*-genetic variants located within a ±1 Mb region and occupied by transcription factors^35^. Elastic-Net is used as the predictive model:

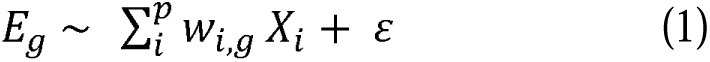

Where *Eg* represents the *g*-th gene expression (AS, APA or cell type-specific gene expression) level;*Xi* denotes the genotype vector of the *i*-th variant (*Xi*=0, 1, or 2) and *p* is the total count of variants within the *cis*-region. The term *w_i,g_* represents the effect size of *i*-th variant for *g*-th gene expression, and *ɛ* is the residual with variance σ^2^*_e_*. An elastic-net penalty, combining LASSO (*L_1_*) and ridge (*L_2_*) regularizers, is applied to optimize (*w_i,g_*) for genetic variants. The Elastic-Net mixing parameter was fixed at *α*=0.5, providing an equal balance between *L_1_* and *L_2_* penalties. The regularization parameter *λ* was selected using three repeated 10-fold cross-validation runs with mean squared error as the loss function. A common sequence of candidate *λ* value was evaluated across the three runs, and the optimal *λ* was defined as the value that minimized the mean cross-validated error across all runs. The final model was fitted using the selected *λ*, and SNPs with nonzero regression coefficients were retained as predictors and used to construct the model weights. Model performance was assessed by calculating the squared correlation (*r^2^*) between predicted and measured gene expression (or AS, APA or cell type-specific gene expression) and average cross-validated *R^2^*(CV *R^2^*) across all cross-validation folds. Models with average cross-validated *R^2>^*0.01 and squared correlation *r^2^*>0.01 are considered as predictable models for downstream meta-analyses.

#### Association analysis

Under our sTF-TWAS framework, we applied our ancestry-specific prediction models (gene expression, AS, APA and cell type-specific gene expression) into GWAS summary statistics from European (49,022 IBD cases and 1,293,489 controls) and East Asian populations (14,393 cases and 15,456 controls), respectively, using the

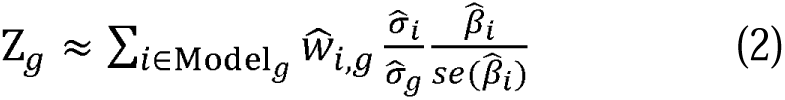

where, the Z*_g_* estimates the association between genetic prediction expression (or AS, APA, cell type-specific gene expression) and IBD risk;*ŵ_i,g_* is the weight of *i*-th genetic variant for predicting *g*-th gene expression.*β̂_i_* and se (*β̂_i_*) are the GWAS-reported regression coefficients estimate and its SE for *i*-th variant; and *σ̂_i_* and *σ̂_g_* are the estimated variances of *i*-th variant and the predicted expression of *g*-th gene (or AS, APA, cell type-specific gene expression). We conducted TWAS, AS-WAS, APA-WAS, and cell type-specific TWAS separately in European and East Asian populations. To integrate gene-level association signals across ancestries and enhance statistical power, we performed meta-analysis using the Aggregated Cauchy Association Test (ACAT). Parallel analyses were conducted for CD and UC, respectively. Statistical significance for TWAS, AS-WAS, APA-WAS, or cell type-specific TWAS associations was determined using analysis-specific Bonferroni correction, with the raw *P* threshold calculated as 0.05/*n*, where n represents the number of bulk genes expression models (*n* = 15,307), AS models (*n* = 20,914), APA models (*n* = 4,380), or cell type-specific gene expression models (*n* = 6,230 for ABS, 772 for GOB and 5,619 for STM) tested in each corresponding disease and ancestry specific analysis.

#### Fine-mapping analysis of identified risk genes

We performed fine-mapping analysis to nominate putative candidate risk genes using FOCUS^10^, for TWAS, AS-WAS, APA-WAS, or cell-type specific TWAS, respectively. The primary association signal for each gene in TWAS (or other molecular trait) was first identified based on ancestry-specific results from either the European or East Asian population. LD reference panels from the corresponding populations in the 1KGP^55^ were used. Fine-mapping was then conducted separately for each ancestry-driven gene set using FOCUS-defined genomic regions and ancestry-matched LD panels. Each gene was assigned a PIP, representing the likelihood of being causal for the observed TWAS signal. Finally, we prioritized candidate genes using two complementary strategies. For genomic regions analyzed by FOCUS, genes with a posterior inclusion probability (PIP) > 0.5 were retained as statistically prioritized candidates, representing majority posterior support rather than definitive evidence of causality. For genomic regions not evaluated by FOCUS, we retained the gene with the most significant TWAS association within a ±1 Mb window around the lead variant. For a detailed description of additional data quality control procedures, data processing, and analytical methods, please refer to the **Supplementary Methods**.

### Evaluating prioritized candidate risk genes through colocalization and SMR analyses

We evaluated their evidence using colocalization analysis for the genes identified by TWAS, AS-WAS, APA-WAS, or cell-type specific TWAS. Colocalization analysis was applied to GWAS and xQTL summary statistics within the ±1 Mb *cis* windows, with strong colocalization defined as posterior probability for a shared causal variant PP.H4 > 0.8. In addition, we evaluated these genes using SMR^57^. Analyses were conducted within the disease, ancestry, and molecular-QTL panel that generated the original association. For each gene-panel, the strongest *cis*-xQTL within ±1 Mb of the transcription start site for variants that was also present in the corresponding GWAS was used as the SMR target SNP. Results were considered supported when they passed the analysis-specific Bonferroni threshold (0.05/*n*), where n is the number of tests performed within each disease-, ancestry-, and xQTL-specific analysis.

Additional methodological details are provided in the **Supplementary Methods**, including sLDSC, pathway enrichment analysis, differential expression analyses on single cell and spatial transcriptomics data, and therapeutic target prioritization.

## DATA AVAILABILITY

The GWAS summary statistics generated from the meta-analysis conducted in this study have been deposited on GitHub, https://github.com/XingyiGuo/IBD_genetics, and also available in the GWAS Catalog (GCP ID: GCP001819). All GWAS summary statistics included in the current study are publicly available. The VA MVP^30^ summary statistics used in this study are available at NIH Genotypes and Phenotypes (dbGaP) under accession number phs002453.v1.p, https://ftp.ncbi.nlm.nih.gov/dbgap/studies/phs002453/analyses/. FinnGen^31^ summary statistics used in this study are available at https://finngen.gitbook.io/documentation/data-download. The Pan-UK Biobank GWAS summary statistics used in this study are available at https://pan.ukbb.broadinstitute.org^32^. The IIBDGC European GWAS meta-analysis^6^ used in this study are available at ftp://ftp.sanger.ac.uk/pub/project/humgen/summary_statistics/human/2016-11-07/. The East Asian GWAS summary statistics used in this study are available at IIBDGC website: https://www.ibdgenetics.org^7^. The RNA-sequencing data and genotype of East Asian ancestry populations from the Asia Colorectal Cancer Consortium used in the study are available at NIH dbGaP under accession number phs002813.v1.p1^35^, https://www.ncbi.nlm.nih.gov/projects/gap/cgi-bin/study.cgi?study_id=phs002812.v1.p1. The data from the GTEx (version 8) used in this study are available through dbGaP under accession number phs000424.v8.p2^33, 34^, https://www.ncbi.nlm.nih.gov/projects/gap/cgi-bin/study.cgi?study_id=phs000424.v8.p2. Transcriptome and genotype data, along with sample covariates, from the BarcUVa-Seq project are available through dbGaP under accession number phs003338.v1.p1^33^, https://www.ncbi.nlm.nih.gov/projects/gap/cgi-bin/study.cgi?study_id=phs003338.v1.p1. Single-cell RNA-sequencing datasets for deconvolution of bulk RNA-seq were obtained from the Colon Molecular Atlas Project (COLON MAP) through the National Cancer Institute-funded Human Tumor Atlas Network (HTAN) Data Portal: HTA11^56^, https://humantumoratlas.org/center/hta11. Single-cell RNA-sequencing datasets used for differential expression analysis are available at the GEO database under accession number GSE266546^27^, https://www.ncbi.nlm.nih.gov/geo/query/acc.cgi?acc=GSE266546. The spatial transcriptomics dataset analyzed in this study was derived from Li et al., Nat Commun 2024^27^. The analysis resources are available at: https://github.com/XingyiGuo/IBD_genetics. LD Scores^58^ from 1000 Genomes is available at https://alkesgroup.broadinstitute.org/LDSCORE. The 1000 Genomes Project reference panel data^55^ (GRCh38) is available at: https://ftp.1000genomes.ebi.ac.uk/vol1/ftp/. Drug-protein data were retrieved from publicly available databases, including ChEMBL^40^: https://www.ebi.ac.uk/chembl/; Therapeutic Target Database^41^: https://db.idrblab.net/ttd/; DrugBank^39^: https://go.drugbank.com/. DisGeNET^42^ database: http://www.disgenet.org. iLINCS database^43^: https://www.ilincs.org/ilincs/. L1000 Connectivity Map^59^: https://www.broadinstitute.org/connectivity-map-cmap. The single-cell eQTL summary statistics from OneK1K^25^ is available at https://onek1k.org. Kyoto Encyclopedia of Genes and Genomes (KEGG)^60^: https://www.genome.jp/kegg/. Reactome^61^: https://reactome.org. Immunology Database and Analysis Portal (ImmPort)^62^: https://immport.org/shared/home.

## CODE AVAILABILITY

The code used our study is available at GitHub: https://github.com/XingyiGuo/IBD_genetics and Zenodo (DOI: 22905952).

## ACKNOWLEDGEMENTS

We acknowledge the significant genetic resource contributions from previous IBD GWASs, VA MVP, FinnGen study, UKB, UKIBDGC and IIBDGC.

## AUTHOR CONTRIBUTIONS STATEMENT

X.G. conceived and designed the study. L.L. and Q.Li. performed the primary data analysis and drafted the manuscript. C.L. contributed to single-cell differential expression analysis and interpretation. G.D., Y.C., R.T., Q.Liu., W.W., X.S., K.W., D.S., W.Z., Z.Y., and K.L contributed to manuscript drafting. Q.Liu. and K.L. contributed to data collection. All authors contributed to results interpretation, manuscript revision, and approved the final version for submission.

