## Supplementary Information for "Multi-ancestry genome-wide and transcriptome-wide association analyses identify new risk loci and genes for inflammatory bowel disease"

**Supplementary Methods:**

**Quality control and association analysis in previous individual GWASs**

In VA Million Veteran Program, the ancestry assessment was followed by genetically inferred ancestry (GIA). Variants with an imputation quality r^2^ > 0.3 and a minor allele count (MAC) > 20 within the relevant (harmonized ancestry and race/ethnicity) HARE-defined ancestry group were included in the analysis^1^. Analyses were adjusted for age, sex, and 10 ancestry-specific genetic principal components (PCs). In FinnGen^2^, association tests were performed for variants with a minimum allele count of 5 among each phenotype’s cases and controls, with sex, age, 10 PCs, FinnGen chip version 1 or 2, and legacy genotyping batch included as covariates. Among the Pan-UK Biobank (UKB)^3^, variants with Imputed Information (INFO) scores over 0.8 and an allele count of ≥ 20 were retained. Sex and age along with the first 10 PCs were included in the GWAS modelling. In the inflammatory bowel disease (IBD) Genetics Consortium genome-wide association study (GWAS) meta, variants with minor allele frequency (MAF) < 0.1% and INFO scores < 0.4 were removed. The sites with strong evidence for heterogeneity (*I*^2^ > 0.9) were discarded in the output of the fixed-effects meta-analysis. In addition, genome-wide significant variants for which the meta-analysis *P*-value was not lower than all the cohort-specific *P*-values were removed. In the East Asian GWAS, variants with an imputation quality r^2^ > 0.6 and MAF > 0.001 were retained in the analysis, adjusting for the first 10 PCs. Details about the QC and association analysis process can be found on the original studies in **Data Availability**.

**Quality control and meta-analysis of GWAS summary statistics**

We first used the LiftOver tool^4^ on the University of California Santa Cruz (UCSC) Genome Browser to convert GWAS summary statistics on GRCh 37 to GRCh 38. Single Nucleotide Polymorphisms (SNPs) that are not able to be converted successfully were dropped from the analysis. For each GWAS summary statistics, if duplicate SNPs were found, the SNP with the smallest *P*-value was retained in the meta-analysis. In the Pan-UKB cohort, only the alternate allele frequency in cases and controls was provided, so the MAF for them was first calculated, and the MAF for the overall population was approximated by averaging the MAF among cases and controls. If the allele frequency or MAF for all populations, neither case nor controls, was not provided in the individual GWAS, we utilized 1000 Genomes Project (1KGP) on GRCh38 European reference panel^5^ to map MAF in the datasets. For meta-analysis, we only included variants on autosomes. Genetic variants with MAF > 0.01 remained for downstream analysis.

We conducted an inverse variance weighted meta-analysis of existing GWAS summary statistics in European populations, including data from MVP, FinnGen (R12), Pan-UKB, and IIBDGC GWAS meta-analysis of IBD using METAL (release 2011-03-25)^6^. Cochran’s *Q* Statistic and *I*^2^ statistics were also calculated using METAL. In the multi-ancestry meta-analysis, we further combined data from East Asians using the same model. Genomic control was applied to each individual GWAS in METAL and then applied to the GWAS meta-analyzed summary statistics according to the METAL document (https://genome.sph.umich.edu/wiki/Metal_Documentation).

Genomic inflation factors (*λ*) and quantile–quantile (Q-Q) plot were utilized to evaluate the genomic inflation of GWAS. If *λ* close to 1, it suggests that genomic inflations were well-controlled. We next applied linkage disequilibrium (LD) score regression (LDSC) regression^7^ by regressing the *χ*² statistics of each SNP from GWAS summary statistics on its corresponding LD score. The pre-computed LD scores were obtained from <https://alkesgroup.broadinstitute.org/LDSCORE>. LDSC intercepts were also considered when evaluating genomic inflation. If the intercepts from LDSC show values close to 1, then the deviation of Q-Q plot may be due to polygenic effects instead of population stratification. To evaluate such effects, we further plotted the Q-Q plot by removing the loci that are located within 300 Kb of previously identified GWAS loci. If the Q-Q plot of removing previously identified loci shows minimal deviation, then the confounding of population stratification was well-controlled.

**Identification of novel loci through LD clumping and conditional analysis**

The novel loci were defined following the previous GWASs of IBD^8, 9^. A locus was defined as known if it overlapped a previously reported IBD locus or if its lead variant was located within 500 kb of a previously reported genome-wide significant IBD-associated variant. We first created a reference risk SNP list for each trait (Crohn’s disease (CD), ulcerative colitis (UC), or IBD) from previous individual GWASs or IBD consortia that were included in the current meta-analysis^1, 2, 8, 9, 10^ by selecting the SNPs that reached the genome-wide significant threshold (*P* < 5×10^-8^). If an SNP was reported in multiple GWASs, the one with the lower *P*-value was retained. For the reference SNP list of IBD, variants previously reported as CD- or UC- associated were also included in the list and considered as IBD-associated, while a known CD-associated SNP was not considered as associated with UC, or vice versa^8^. Next, we selected the genome-wide significant (*P* < 5×10^-8^) SNPs from our meta-analysis and removed the variants on the reference SNP list. To ensure the putatively identified genome-wide significant SNPs in our meta-analysis were not due to LD with previously reported variants, we further checked whether they are at least 500 Kb away from already known SNPs. If yes, the SNP was not considered novel and was removed. For the remaining genome-wide significant SNPs, we performed a LD clumping with an LD window of 500 Kb and r^2^ threshold of 0.6 using the 1KGP European reference panel for European-specific meta-analysis to identify the novel genetic loci. If the genetic loci after LD clumping overlapped with other loci within 500 Kb, we then merged the loci and kept the variants with the strongest association (the lowest *P*-value) as the lead variants for the merged genetic loci. Clumping was performed using PLINK 1.9^11^. In addition, the retained SNPs were further cross-checked with GWAS loci identified from the latest GWASs of IBD^9^ to confirm that they are novel.

For the multi-ancestry meta-analysis, the 1KGP European reference panel and 1KGP East Asian reference panel were used for LD clumping, respectively. LD clumping results from the European and East Asian panel were merged, and the lead variants with the lowest *P* within 500 Kb were kept. To identify additional novel loci from European-specific meta-analysis, we merged the overlapping loci from European-specific and multi-ancestry meta-analysis within 500 Kb and then compared the *P*-values for the lead variants in the overlapping loci. Loci where lead variants from multi-ancestry meta-analysis with the lower *P*-values were considered as the additional novel loci from multi-ancestry meta-analysis.

To ensure that putatively novel signals were not due to long-range LD with previously reported loci, we conducted conditional analysis^12^ using the summary statistics of our GWAS meta-analysis for the lead variants in loci that are within 1Mb of a known locus. The candidate SNP is conditioned on the nearest previously reported IBD lead variant. Signals with *P*-value < 5×10^-6^ after conditioning on the nearest known lead variant were considered conditionally independent, whereas loci with *P*-value > 5×10^-6^ or non-estimable conditional associations were not considered as independent novel loci.

Novel loci were defined as those whose lead variants were located more than 1 Mb from any previously reported IBD susceptibility locus or remained independently (*P* < 5 × 10^−6^) associated after conditioning on the nearest previously reported lead variant located within 500 Kb to 1Mb interval. This definition was applied across IBD-related traits. Specifically, variants previously reported as CD- or UC-associated were considered known IBD-associated variants when defining novel loci for overall IBD. In contrast, variants previously reported only for IBD were not considered known CD- or UC-associated variants in subtype-specific analyses; similarly, known CD-associated SNPs were not considered UC-associated, and vice versa. Thus, loci previously reported for one IBD subtype but showing an independent association with another subtype in the present study were retained as novel subtype-associated loci.

**Genotype data processing**

For the BarcUVa-Seq project^13^, 400,000 SNPs from was imputed using TOPMed (Version 2) with European ancestry as the reference panel. We then excluded variants with an imputation quality R^2^ < 0.3, missingness rate > 0.1, a MAF < 0.01, or significant deviations from Hardy-Weinberg equilibrium (*P* < 10^-6^). The same process was applied to genotype data from East Asian data, which was imputed using TOPMed with Asian ancestry as reference panel^14^. In addition, we obtained the whole genome sequencing (WGS) data from Genotype-Tissue Expression (GTEx) project and dbGaP (phs000424.v8.p2)^13, 15^. A comprehensive outline of WGS variant quality control procedures is available from the GTEx consortium^15^.

**RNA-seq data processing and gene expression, AS and APA quantification**

We developed ancestry-specific transcriptome-wide association study (TWAS) models using three bulk gene expression datasets: 707 European-ancestry samples, including 423 from the BarcUVa-Seq project^13^ and 284 from the GTEx project^13, 15^, along with 364 Asian-ancestry samples from our previous study^16^. Gene-level expression was quantified using RNA-SeQC, aligning with the annotation used in ^15^ as described in our previous study^13^. In general, we applied quantile and inverse normalization to the RNA-seq transcript per million (TPM) data. Next, we adjusted potential confounding factors through a probabilistic estimation of expression residuals (PEER) analysis, following the GTEx recommendations for determining the number of PEER factors. We quantified AS level from RNA-seq alignments using LeafCutter^17^ and RegTools^17^, following the methods descried in our previous study^13^. RegTools was used to extract exon-exon splice junctions from the BAM files, after which LeafCutter grouped overlapping introns into clusters and calculated the proportion of junction-spanning reads supporting each alternatively excised intron. The resulting intron-excision ratios were used as the molecular phenotypes for constructing AS prediction models. For each event, models were trained using *cis* variants flanking the splice site, with model parameters selected by tenfold cross-validation^13, 17^. The level of APA was assessed using the percentage of PDUI, as estimated with DaPars v2.0^18^, in accordance with our prior research^19^.

For downstream gene expression, AS and APA prediction model building, we adjusted these measurements for potential confounding factors, including PEER factors (60 for BarcUVa-Seq and Asian-ancestry data, 45 for GTEx), as well as age, sex, and the first five genotype PCs.

**Cell type-specific gene expression quantification (****deconvolution bulk RNA-seq based on the reference single-cell data.)**

We obtained single-cell RNA sequencing (scRNA-seq) data for 33,806 epithelial cells from normal colon tissue using the Colon Molecular Atlas Project^20^. These epithelial cells are clustered into seven clusters: absorptive cells (ABS), crypt top cells (CT), enteroendocrine cells (EE), goblet cells (GOB), stem cells (STM), transit amplifying cells (TAC), and tuft cells (TUF). We applied quality control to select high quality cells: nUMI > 500, nGene > 250, log_10_GenesPerUMI > 0.8, and mitoRatio < 0.2. Following the CIBERSORTx^21^ pipeline, we constructed a signature gene expression matrix for each cell type and estimated each cell type fraction for bulk RNA-seq data from 707 European and 364 East Asian individuals. Among these analyzed cell types, ABS cells exhibited the highest median proportion (49%), followed by STM-like cells (29%), GOB cells (3%), and other cell types (< 1%). We kept the top three most abundant cell types (ABS, STM, and GOB) and performed deconvolution of bulk RNA-seq data using their respective signature matrices to derive cell type-specific gene expression profiles. The downstream processing and normalization of cell type-specific gene expression data were consistent with those used for bulk gene expression prediction. Model building for cell type-specific expression also incorporated the recommended PEER factors, along with age, sex, and the first five genotype PCs.

**Immune cell-specific *cis*-eQTL analysis using the OneK1K resource**

In addition to xQTL analyses in normal colon tissue and epithelial cell types, we incorporated immune cell-specific *cis*-eQTL data from the OneK1K single-cell RNA-seq resource to further evaluate immune cell-specific regulatory effects. We extracted novel GWAS lead variants from our ancestry- and disease-specific analyses and mapped these variants to the OneK1K immune cell-specific *cis*-eQTL database^22^. For each overlapping lead variant, we identified the corresponding target gene and immune cell type in which the variant exhibited significant *cis*-eQTL effects. The identified immune cell-specific eQTLs were used as complementary functional evidence to support the prioritization of candidate target genes and to provide additional insight into the immune cell types through which genetic risk variants may influence IBD susceptibility.

**Stratified LDSC regression analysis**

To identify the most disease-relevant cell types in colon tissue for IBD and its subtypes, we performed a stratified LDSC (sLDSC) by using the single-cell normalized gene expression matrix and GWAS summary statistics of IBD. The scRNA-seq gene expression data on terminal ileum (TI) and ascending colon (AC) tissues were obtained from 202,359 cells from 170 specimens across non-IBD controls (N = 18) and active/inactive CD patients (N = 65) from the National Center for Biotechnology Information Gene Expression Omnibus (GEO)^23^. The cells were classified into 16 cell types within the epithelial compartment: early enterocytes, intermediate enterocyte, mature enterocyte, early colonocyte, intermediate colonocytes cells, mature colonocyte, early GOB, mature GOB, GOB proliferating cells, cycling TA cells, BEST4/OTOP2 cells, tuft cells, enteroendocrine cells (EEC), Paneth cells, STM cells, and LND cells (annotated by high expression of *LCN2*, *NOS2*, and *DUOX2*)^23^. Among them, enterocytes and colonocytes were merged as ABS cells. ABS, GOB, and STM were considered as major cell types for IBD and included in the follow-up analysis. Using Seurat v5^24^, we generated a pseudo bulk gene expression matrix for each of the 83 subjects (18 non-IBD controls and 65 CD patients). Pseudo bulk aggregation was used to sum raw counts across all cells of a given type within each subject, reducing the influence of outlier cells and producing more robust, cell type-specific expression profiles^20^. Genes were then ranked by *t*-statistic within each cell type, and the top 10% were defined as specifically expressed genes^25^. Finally, we applied sLDSC on specifically expressed genes to estimate partitioned heritability and identify cell types significantly enriched for IBD risk.

**Pathway enrichment analysis and function categorization**

To explore the biological functions and pathways enriched in the gene list, we performed gene set enrichment analysis using the Enrichr web-based tool (https://maayanlab.cloud/Enrichr/). Enrichr integrates multiple curated databases^26, 27, 28^, enabling comprehensive functional annotation and pathway enrichment analysis. Enrichr uses a combined score metric that incorporates the *P*-value from the Fisher exact test and the z-score of the deviation from the expected rank to rank enriched terms. Significantly enriched pathways were identified based on false discovery rate (FDR)-corrected *P* < 0.05. To contextualize these genes within known biological processes, we mapped the identified genes to established IBD-related pathways. Pathway definitions were obtained from curated databases, including Kyoto Encyclopedia of Genes and Genomes (KEGG)^29^ (https://www.genome.jp/kegg/), Reactome (https://reactome.org/)^30^, and the Immunology Database and Analysis Portal (ImmPort)^31^ (https://immport.org/shared/home). Additionally, we referenced literature-supported functional experiments that validate gene involvement in IBD pathogenesis. We further annotated genes with supporting evidence from functional studies, including expression profiling, knockout or knockdown models, and immunological assays, to highlight genes with demonstrated mechanistic roles in IBD.

**Differential expression analysis using single-cell RNA-sequencing and spatial transcriptomics**

Pseudo-bulk aggregation with summing raw counts for each cell type from 18 non-IBD controls and 65 CD patients^23^ was used, as described in the preceding section. We performed differential expression analysis for each cell type between non-IBD controls and active/inactive CD subjects using DESeq2 with default parameters. Genes with FDR-corrected *P* < 0.05 were considered as significantly differentially expressed genes. If a significant gene identified by TWAS and xQTL also showed an average log_2_ fold change in the same direction in the single-cell analysis and had an FDR-corrected *P*-value < 0.05, it was considered to have a consistent direction of association with the TWAS and xQTL results.

Differential expression analysis was also performed in spatial transcriptomic data obtained from CD samples from 10X Genomics. Four CD samples from the above samples were selected because of their relatively high proportions of LND cells^23^. Among them, two samples were with active terminal ileum (TI) disease (GCA092 and GCA033), while the other two were with active ascending colon (AC) disease (GCA089 and GCA099)^23^. Data normalization and scaling were performed in R using SCTransform and ScaleData functions, respectively. To identify spatially variable genes, we used FindVariableFeatures function to select the most informative genes for downstream analysis. Principal component analysis (PCA) was used to reduce dimensionality using RunPCA function, and the top 10 PCs were used for clustering with FindClusters function at a resolution of 0.8. FindNeighbors function was used to compute a nearest-neighbor graph based on PCA. For differential expression analysis, the FindAllMarkers function was used to compare the gene expression profile between one cluster and all others within the same sample, with the minimum percentage expression threshold of 0.25 and log fold-change threshold of 0.25. *P*-values were adjusted using FDR correction, and corrected *P* < 0.05 were considered as statistically significant genes. Uniform Manifold Approximation and Projection (UMAP) were performed for visualization.

Cell type annotation for each cluster was conducted using GPTCelltype^32^ by selecting the top positively expressed feature genes. Clustering visualization was presented in **Supplementary Figure 5**.

**Characterization of drugs associated with prioritized candidate genes**

To identify potential therapeutic drug candidates, we assembled a comprehensive set of drug-target pairs from four drug databases: DrugBank^33^, ChEMBL^33^, and the Therapeutic Target Database (TTD)^34^. DrugBank is a curated database containing over 500,000 drugs and thousands of bioentities (proteins, genes, and other cellular components). ChEMBL provides detailed information on more than 2 million bioactivity assays for bioactive molecules with drug-like properties, covering over 2 million compounds. TTD offers comprehensive information on thousands of drug targets, including 426 approved, 1,014 in clinical trials, 212 in preclinical or patented stages, and 1,479 reported in the literature. Drugs and their paired genes were comprehensively downloaded from these three databases. We subsequently linked these drugs to our identified prioritized candidate risk genes.

To assess the disease specificity of the potential druggable genes, we downloaded genes associated with IBD, UC, and CD from DisGeNET^35^ using the corresponding Concept Unique Identifiers: C0021390 for IBD, C0009324 for UC, and C0156147 for CD of the large bowel.

To further explore therapeutic potential, we searched for compounds capable of reversing gene expression signatures using the iLINCS database^36^. iLINCS provides transcriptomic and proteomic perturbation signatures describing gene expression changes induced by chemical or genetic perturbations. We downloaded chemical perturbation (CP) signatures derived from large intestine–related cell lines, each containing average *Z* scores and associated *P*-values for 978 landmark genes from the L1000 Connectivity Map^37^. Following the approach of Wu et al.^38^, we computed Pearson correlations ($r$) between bulk tissue *Z* scores from each dataset and averaged perturbation *Z* scores for the top 50 most upregulated and downregulated genes. Compounds exhibiting significant negative correlations (*P* < 0.05) were prioritized as potential candidates capable of reversing disease-associated transcriptional profiles.

**Supplementary Figures**

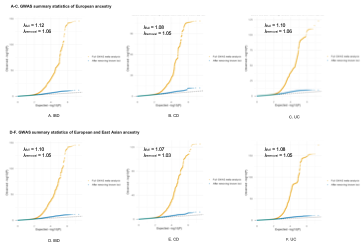

**Supplementary Figure 1. Q-Q plots of GWAS meta-analysis before and after removing variants within ±300 Kb of known IBD loci.** Q-Q plots compare observed and expected -log_10_(*P*) values for GWAS meta-analysis results of inflammatory bowel disease (IBD), Crohn’s disease (CD), and ulcerative colitis (UC). Panels A-C show the European meta-analysis, and panels E-G show the combined European and East Asian meta-analysis. Orange points represent the full GWAS meta-analysis results, while blue points represent results after removing variants within 300 Kb of previously reported genome-wide significant loci. Genomic inflation factors (*λ*) are shown for the full and locus-removed analyses, demonstrating reduced inflation after removal of established association regions.

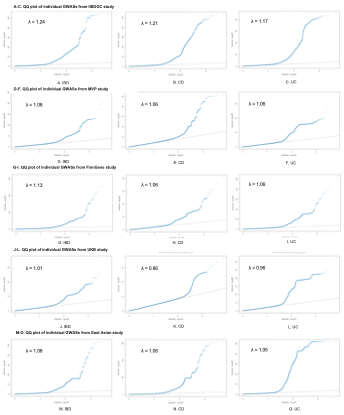

**Supplementary Figure 2. Q-Q plots of** **each contributing GWAS datasets.** Q-Q plots compare observed and expected -log_10_(*P*) values for GWAS meta-analysis results of inflammatory bowel disease (IBD), Crohn’s disease (CD), and ulcerative colitis (UC). Panels A-C show the European meta-analysis, and panels E-G show the combined European and East Asian meta-analysis. Orange points represent the full GWAS meta-analysis results, while blue points represent results after removing variants within 300 Kb of previously reported genome-wide significant loci. Genomic inflation factors (*λ*) are shown for the full and locus-removed analyses, demonstrating reduced inflation after removal of established association regions.

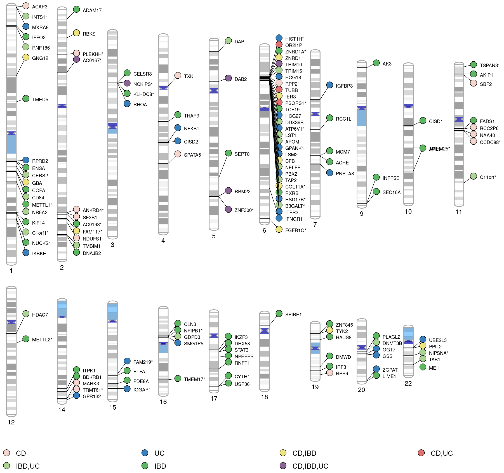

**Supplementary Figure 3. Novel prioritized candidate risk genes identified across IBD, CD, and UC from cell type-specific TWAS analyses.** 134 novel prioritized candidate risk genes identified by TWAS, APA-WAS, and AS-WAS are displayed according to their chromosomal positions. Genes are grouped according to their phenotype associations: "IBD" (unique to inflammatory bowel disease), "CD" (unique to Crohn’s disease), "UC" (unique to ulcerative colitis), "CD,IBD" (shared by CD and IBD), "CD,UC" (shared by CD and UC), "IBD,UC" (shared by IBD and UC), and "CD,IBD,UC" (shared by all three phenotypes). Gene names are truncated with remaining characters replaced by an asterisk (*) due to space constraints. Associations were evaluated using two-sided Wald *z*-tests implemented in sTF-TWAS. Statistical significance was determined separately for each analysis using a Bonferroni-corrected threshold of *P*/*n* where *n* was the number of models tested in the corresponding disease- and ancestry-specific analysis.

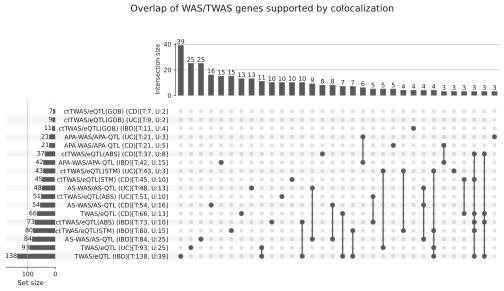

**Supplementary Figure 4.** **Unique and shared prioritized candidate risk genes identified across IBD, CD, and UC.** The plots summarize the overlap among prioritized candidate risk genes identified through TWAS, AS-WAS, APA-WAS, cell-type-specific TWAS and xQTL mapping within GWAS risk loci for IBD, CD, and UC. Horizontal bars show the total number of genes identified by each analysis. Vertical bars show the number of genes in each indicated intersection. A single filled circle denotes genes identified exclusively by the corresponding analysis, whereas filled circles connected by a black line denote genes shared exclusively among the indicated analyses and not identified by the remaining analyses. The figure summarizes overlaps among prioritized genes; no additional statistical test was performed.

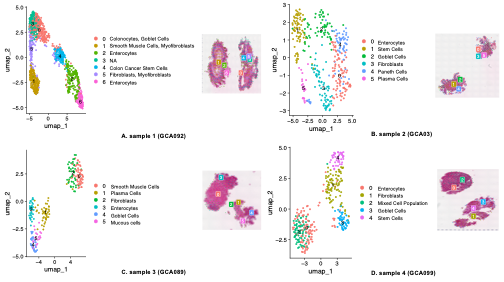

**Supplementary Figure 5**. **Cell-type annotation of spatial transcriptomic profiles from four colon tissue samples.** UMAP embeddings and corresponding spatial maps show the cell populations identified in (A) GCA092, (B) GCA03, (C) GCA089, and (D) GCA099. In each panel, points in the UMAP represent spatial transcriptomic spots and are colored according to their assigned cell type; the corresponding tissue image shows the spatial distribution of these annotations. Cell identities were assigned using GPTCelltype^32^ by selecting the top positively expressed feature genes. Cluster numbers correspond to the cell populations indicated in each panel’s key.
